# Chagas disease: lack of knowledge as a risk factor

**DOI:** 10.1101/2024.04.09.24301992

**Authors:** Nicolás Mario Mas D’Alessandro, Hernán Daniel Blaisten, Cristian José Ferrufino, Julián Matías Fernández Boccazzi

**Affiliations:** MetroHealth; Fundacion Favaloro

## Abstract

Chagas disease presents not merely as a medical concern but as a multifaceted socio-environmental health issue demanding diverse approaches. In Argentina, an estimated two million individuals are afflicted with Chagas disease, with 600,000 exhibiting clinical manifestations. This study endeavors to gauge the populace’s knowledge levels in Añatuya, situated northeast of Santiago del Estero, and to compare comprehension levels between Chagas disease patients and non-patients. Employing a cross-sectional epidemiological design, demographic, socio-economic, and medical data were gathered through surveys from individuals aged 18 and above attending local healthcare facilities. Analysis revealed that among 200 patients, 12% exhibited low, 66% intermediate, and 22% high levels of elementary knowledge regarding Chagas disease, nearing optimal understanding for an endemic region. However, statistically significant differences (p < 0.05) in knowledge were observed between Chagas and non-Chagas patients in only three out of ten elementary knowledge questions. Moreover, housing quality, a pivotal risk factor for Triatoma Infestans presence, indicated that 78% resided in improved dwellings, yet 22% lived in high-risk housing, reflecting persistent precarious living conditions susceptible to infestation. Enhanced comprehension of Chagas disease fundamentals and associated risk factors could empower endemic area residents towards proactive well-being practices, advocating for improved living standards and disease management strategies.

## Introduction

Chagas is a serious public health problem in Argentina and throughout Latin America. It is an endemic disease in this territory, distributed from Mexico to South America, having a greater prevalence in rural areas of Latin America, although there are vectors and reservoirs even in the south of the United States and with some cases identified in Canada. In these areas mentioned, it is estimated that there are between 8 and 11 million people infected, and it is estimated that 10,000 people die annually from this disease. (1)

According to the WHO (2010), there are 5,742,167 people infected with T. Cruzi in 21 Latin American countries, of which 62.4% (3,581,423) are from countries in the Southern Cone. Argentina, Brazil and Mexico were the 3 countries with the highest estimated number of infected people (1,505,235, 1,156,821, 876,458, respectively), followed by Bolivia (607,186). However, this last country has the highest number of new cases due to vector transmission (8,087), with Argentina being seventh on the continent in terms of this index (1,078). Regarding congenital transmission, our country is in second place with 1,457 new estimated cases annually, a ranking led by Mexico (1,788). (2)

It currently occupies fourth place in importance as a cause of disability, after respiratory diseases, diarrhea and AIDS. In Argentina, it is estimated that there are between one and a half and two million infected people, 600,000 of them with clinical manifestations (3) (4). The importance of this parasitosis lies in its high prevalence, large economic losses due to work incapacity and death. sudden onset of apparently healthy people.

Triatomines proliferate in houses with poor conditions (for example, mud walls and thatched roofs), which is why people living in rural areas in countries where the disease is endemic are exposed to a higher risk of contracting the infection.

Chagas Disease or American Trypanosomiasis is a tropical parasitic zoonosis produced by the flagellated protozoan Trypanosoma cruzi, a single-celled parasite that is transmitted mainly by a blood-sucking insect, popularly called “vinchuca” in our country. The most important species in the Southern Cone is Triatoma infestans, which lives inside the home and peridomicile. This protozoan enters the insect’s digestive tract when it bites a person or more than 100 species of domestic and wild mammals. After the vector sucks blood at night, the parasite actively divides in the insect and is transmitted through its feces, which are deposited in the excoriation or wound on the host’s skin, which may well have been generated by the bite of the vinchuca, orally or ocularly. This route of transmission, called vector, is the most common in the region of our country, although it is not the only one, since it can also occur through other routes: oral (through contaminated food), congenital, through contaminated blood products. (blood transfusion, organ donation), organ transplant by infected donors or laboratory accident. (5)

The clinical manifestations can be diverse: during the acute phase they are generally asymptomatic or oligosymptomatic, although chagoma or, in a minority, the Romaña sign may occur. The indeterminate phase is usually asymptomatic, although fever, anorexia, lymphadenopathy, mild hepatosplenomegaly, and myocarditis may occur. The chronic phase is symptomatic and can appear decades after the initial infection, affecting the nervous system (dementia), digestive system (megacolon, megaesophagus) and the heart. After several years, 30% of those infected will develop heart damage (right bundle branch block or left anterior fascicular block, dilated cardiomyopathy), 6% digestive disorders and 3% disorders of the peripheral nervous system, which can be fatal. because of the cardiac component. Sudden cardiac death is the most common cause of death (55-65% of patients), followed by congestive heart failure and thromboembolism (6). In fact, it is estimated that Argentina is the country with the highest number of Chagasic people with heart disease (376,309 according to the WHO). (7) (2)

Its diagnostic criteria are based on a history compatible with the epidemiology of the disease and microbiological studies (direct and/or indirect parasitological techniques) and its treatment is specific for the parasite or symptomatic of the complications, depending on the phase in which it is found. the patient. The drugs used and provided by the National Chagas Program are Benznidazole and Nifurtimox. (5) (8) (9)

However, Chagas is not just a disease, it is a complex and multidimensional socio-environmental health problem that has different points of approach. It covers four large dimensions: the biomedical (biology of the parasite and the transmitting insect, clinical manifestations of the disease, diagnosis, treatment and transmission route), the epidemiological (characterizes the situation at the population level, the geographical problems, migratory movements, climate change).), the socio-cultural (cultural practices, environmental management, particularities of the rural and urban context, stereotypes and prejudices, discrimination and stigmatization) and politics (public management and decision-making in the health, educational and legislative fields at the local level, regional and global, resource management). Although they are different dimensions, their limits are diffuse, they are all intertwined and play a fundamental role in the perpetuation of the disease (10). It is recognized by the World Health Organization (WHO) as one of the 13 most neglected tropical diseases in the world, and by the Pan American Health Organization as a disease of poverty. (eleven)

Because an effective vaccine has not yet been developed to prevent the disease, strategies for its control try to reduce transmission, mainly vector transmission. These control actions are aimed at the chemical attack of the vectors without taking into account that there are many other risk factors responsible for the persistence of triatomine foci.

Simply spraying houses with insecticides is not a completely effective action to eradicate the disease. It is a trend continued since the development of the National Control Programs in our country (1962) that is insufficient. According to the Ministry of Health, upon approving the National Chagas Plan, the provinces of Chaco, Formosa, Santiago del Estero, San Juan, Mendoza and Córdoba present a re-emergence of vertical transmission of Chagas due to an increase in household infestation and a high seroprevalence in vulnerable groups, the situation being classified as “high risk for vector transmission” (3) (4). According to the WHO, the estimated prevalence of vector infection per 100 inhabitants is the highest on the continent in the areas that cover the Gran Chaco region (belonging to Bolivia, Argentina and Paraguay).

We believe that knowledge on the part of the population and the authorities who make decisions is a key point that fits perfectly within the aforementioned dimensions; it depends on both education and management, the actions of the doctor and the community. This can be an approach that, if given due importance, can be highly beneficial in the fight against the disease. That is why we propose misinformation or ignorance about Chagas disease as a risk factor, since it would not be known how to try to combat it. That is, the level of knowledge of the inhabitants of endemic areas about the disease and its vectors should be one more element for its prevention and control.

The present work corresponds to a population sample from the city of Añatuya, located southeast of the province of Santiago del Estero, capital city of the department of General Taboada, Argentina, whose official population figures are 23,286 inhabitants (INDEC 2010)., although unofficially it is estimated that this year it reaches 40,000 inhabitants. During the diagnosis of the socio-sanitary situation we were able to observe a region with a large number of ranch-type homes, lack of waste collection, contaminated streets due to the absence of sewers, a large number of dogs and chickens in the streets, many of these factors being risk for the presence of the Chagas Disease vector.

For this reason, the objective of this work is to determine the level of knowledge of the inhabitants of the area according to the elementary notions that constitute the optimal level of knowledge (NOC) and the risk factors present in the region. (12)

### General objective

Determine the degree of knowledge about Chagas disease and its vector and the prevalence of its risk factors, in all people over 18 years of age who have attended the Zonal Hospital of the city of Añatuya, the zonal health posts No. 4, 9 and the Villa Mailín health post, covering the area of influence of the hospital (up to towns 100km from the city of Añatuya).

### Main objectives

- Determine the level of knowledge of the inhabitants of the city of Añatuya according to the list of elementary notions that constitute the optimal level of knowledge (NOC) of Chagas Disease and compare if there is a significant difference between those who have the disease and those who do. No.
- Study the socio-cultural and demographic conditions of each patient over 18 years of age surveyed to evaluate whether or not it is related to the risk factors for Chagas Disease.
- Health promotion through health education and disease prevention due to possible transmission routes.
- Make intervention proposals by the health team in the community to contribute to the eradication of the vector

### Secondary objectives

- Contribute to the monitoring of Chagas patients through the development of a database of surveyed patients.
- Detect probable undiagnosed cases of Chagas and refer them to the cardiologist for study.

## Materials and methods

### Type of study

According to the objective of the study, the available means (personnel, information, economic resources, etc.), frequency of the disease or exposure factor to be studied, ethical aspects and other factors, we choose an epidemiological design that correctly adapts to our situation to obtain the desired information in the most accurate way possible. In this way, we carried out an observational study since it seeks to establish the risk factors and only observe characteristics that the population has acquired naturally, without any intervention on the subjects. It is a cross section according to the temporal sequence, since there is no continuity in the time axis of the study, that is, it was located in a single temporal moment. It is mainly descriptive since only the observed data obtained from the population studied are reported. Although it also has an analytical nature, since we compare the responses corresponding to the table of optimal notions of knowledge (NOC Table) between non-chagasic patients and chagasic patients using 2×2 contingency tables and Chi2 as a statistical method. We took a value of p < 0.05 as a statistically significant difference with an α = 5% and CI = 95%. Degrees of freedom = 1.

### Population

The population sample consists of N=200, people over 18 years of age who attended the Zonal Hospital of Añatuya (on-call service, pediatric outpatient clinic, general surgery and medical clinic, maternity, inpatient ward), to the peripheral Health Posts within the City of Añatuya (No. 5 and 9), to the festival of the Lord of Miracles in Villa Mailín, as well as residents of the peripheral neighborhoods to the hospital, between 04/17/2017 and 06/20/2017, to whom we evaluate their knowledge about Chagas Disease.

### Inclusion criteria

Patients will be included in the study if they meet the following criteria:

- Being over 18 years
- Previously accept the completion of the survey
- Be residents within the area of influence of the zonal hospital of the city of Añatuya (100km radius).
- Patients who have attended the on-call services, pediatrics, general surgery, cardiology and medical clinic of the Zonal Hospital of the city of Añatuya, Health Posts No. 4 and 9, and the on-call service of the Villa Mailín health post during on May 24 to 28.

### Exclusion criteria

- Residents outside the area of influence of the Añatuya zonal hospital.
- Be under 18 years of age.

### Human Resources

The research work, as well as the implementation of surveys, were designed by 6th year medical students at Favaloro University: Blaisten, Hernán Daniel; Fernández Boccazzi, Julián Matías; Ferrufino, Cristian José; and Mas D’alessandro, Nicolás Mario, with the collaboration of the cardiologist Dr. Mujica, who allowed our presence in the cardiology consultations and was a reference on the disease; from the pediatrician Dr Seri, María Cristina, who in addition to allowing our presence in the pediatric consultations at the Zonal Hospital to be able to conduct surveys with the children’s parents/companions, guided and accompanied us throughout the work carried out; and all the staff of the Zonal Hospital who contributed their good collaboration and knowledge of the population of the area. The scope of the survey was within the Zonal Hospital, in the health posts and on the streets of the city.

### Materials

200 surveys were designed and printed in Word format. (Annex 1). The survey design consists of a first section, which inquired about affiliation/demographic data: name and surname of the respondent, sex, date of birth, age, nationality, educational level, how many people they live with in their home, occupation. usual, medical coverage or association with a private or public plan and type of housing (home, cohabitants with Chagas, pets, whether or not it is made of material, if it had electricity, drinking water, installed bathroom and waste collection, if you witnessed any sometimes vinchucas inside or outside your home, fumigation). In this way, the number of risk factors present in the sample was obtained.

The second section grouped a core of questions that evaluated the basic knowledge of Chagas Disease and its link with the vinchuca, a large percentage of them being quantitative, dichotomous variables: recognition of the vector and its remains, its habitat, route and method of transmission, organs that affect, among others. To design this section of the survey and thus determine the level of knowledge of the respondents, the total list of “elementary notions” that constitutes the NOC on Chagas Disease was taken as a basis. The NOC is represented by 25 “updated knowledge” referring to the disease and its transmission characteristics that are considered fundamental for all residents in areas where Chagas Disease is endemic, therefore, it fully applies to the region in question. This list of knowledge (Annex 2) arose from the analysis of previous research on the characteristics of the disease and its risk factors (12). It was decided to use 10 questions to representatively encompass the aforementioned list and thus evaluate the respondents and their relationship with the NOC, stratifying them into low, intermediate and high levels of knowledge, according to the bibliography. (13)

In the third and final section, the patients’ medical data were analyzed: periodic health checks and their frequency, relevant diseases, whether they had Chagas analysis ever in their life and what the result was.

If you were a patient with Chagas (having been previously diagnosed or having a positive serology), you were asked how long ago you were diagnosed, if you went to the cardiologist or if you had an electrocardiogram in the last year, symptoms or heart diseases, and, If you received any type of treatment, what medication was administered.

The survey concluded by giving the patient the opportunity to make some type of comment or observation about the survey.

At the same time, the Microsoft Excel spreadsheet corresponding to the coding of the surveys is attached, as well as the Chi2 calculation spreadsheet.

### Procedure

In the first three weeks of the rural Rotation, started on 04/03/2017, a detailed health diagnosis of the population has been carried out, trying to see those pathologies and health problems prevalent in the population that attended the Zonal Hospital of Añatuya. To achieve this, the following sources of information were used: hospitalization and on-call ward medical records, external medical clinic and cardiology offices, demographic data, consultations with the epidemiology and social department provided by the Municipality of the city of Añatuya and health posts, and bibliographic sources. We were notably struck by the number of Chagas patients who frequented the hospital, mainly in the cardiology service. Finally, between the fourth and fifth week, and with the collaboration of Dr. Mujica, it was decided to carry out the work on Chagas Disease from a medical-social perspective. Regarding this aspect, the methodology chosen was to carry out surveys to evaluate the knowledge of the population of the city of Añatuya about Chagas Disease and its link with vinchuca. Once designed and printed, they were implemented during the months of May and June. To this end, with the collaboration of the hospital and its members, we decided to dedicate an external office exclusively to carry out surveys on all patients admitted to the different services. They were not self-administered, but the questions were read to them and doubts about their understanding were answered. The collaboration of all doctors on duty was requested, who redirected their patients to our office once their respective medical appointments had ended. We have the help of Dr. Maria Cristina Seri, a pediatrician, and Dr. Mujica, a cardiologist specializing in Chagas, to be able to attend your consultations and conduct surveys. We also carried out surveys within the regional and rural health posts that Dr. Seri regularly attends (Post number 9 in Barrio Malvinas, Post number 4 of the Terminal station). Simultaneously, we made trips to the different neighborhoods of the city to conduct surveys of neighbors, outside the hospital setting. Taking advantage of our stay during the Festival of the Lord of Miracles that is celebrated in Villa Mailín from June 25 to 28, we continued collecting information in said town.

All people who attended the Añatuya Zonal Hospital were randomly surveyed, with their prior consent.

All information collected from the surveys was coded and centralized in a Microsoft Excel database, attached to the research work delivery folder. It is from the month of May that we begin to carry out this work written in Microsoft Word, with subsequent analysis of the data obtained.

## Results

Results of the first section surveyed: PERSONAL DATA/DEMOGRAPHIC DATA. Of the total number of patients interviewed, 71% (n=142) were female while 29% (n=58) were male.

**Graph 1.**
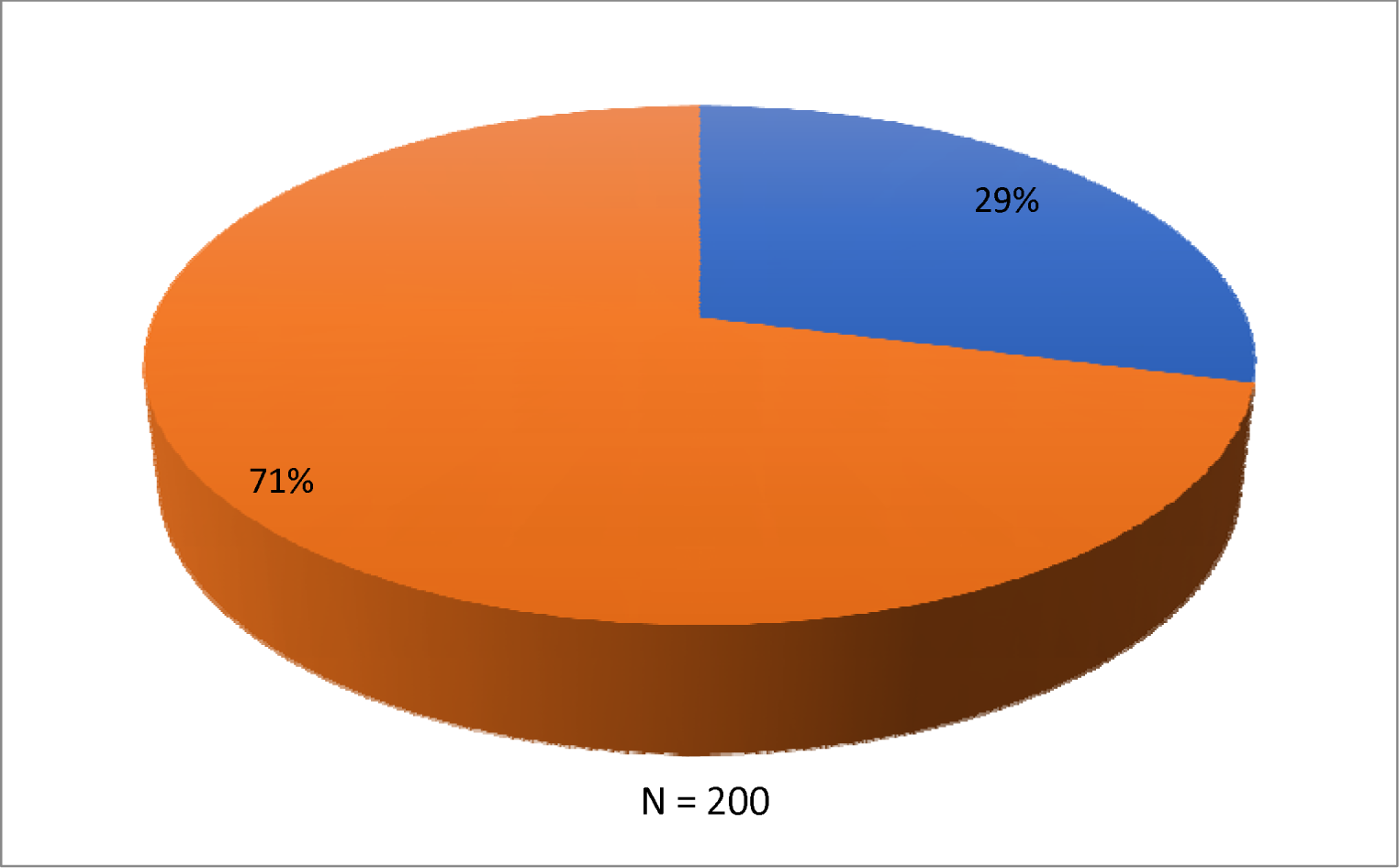
Sex

200 patients aged between 18 and 77 years were evaluated. Of the total patients interviewed, 43% (n=86) are between 18 and 30 years old; 44% (n=87) between 30 and 50 years old and 14% (n=27) over 50 years old.

**Graph 2.**
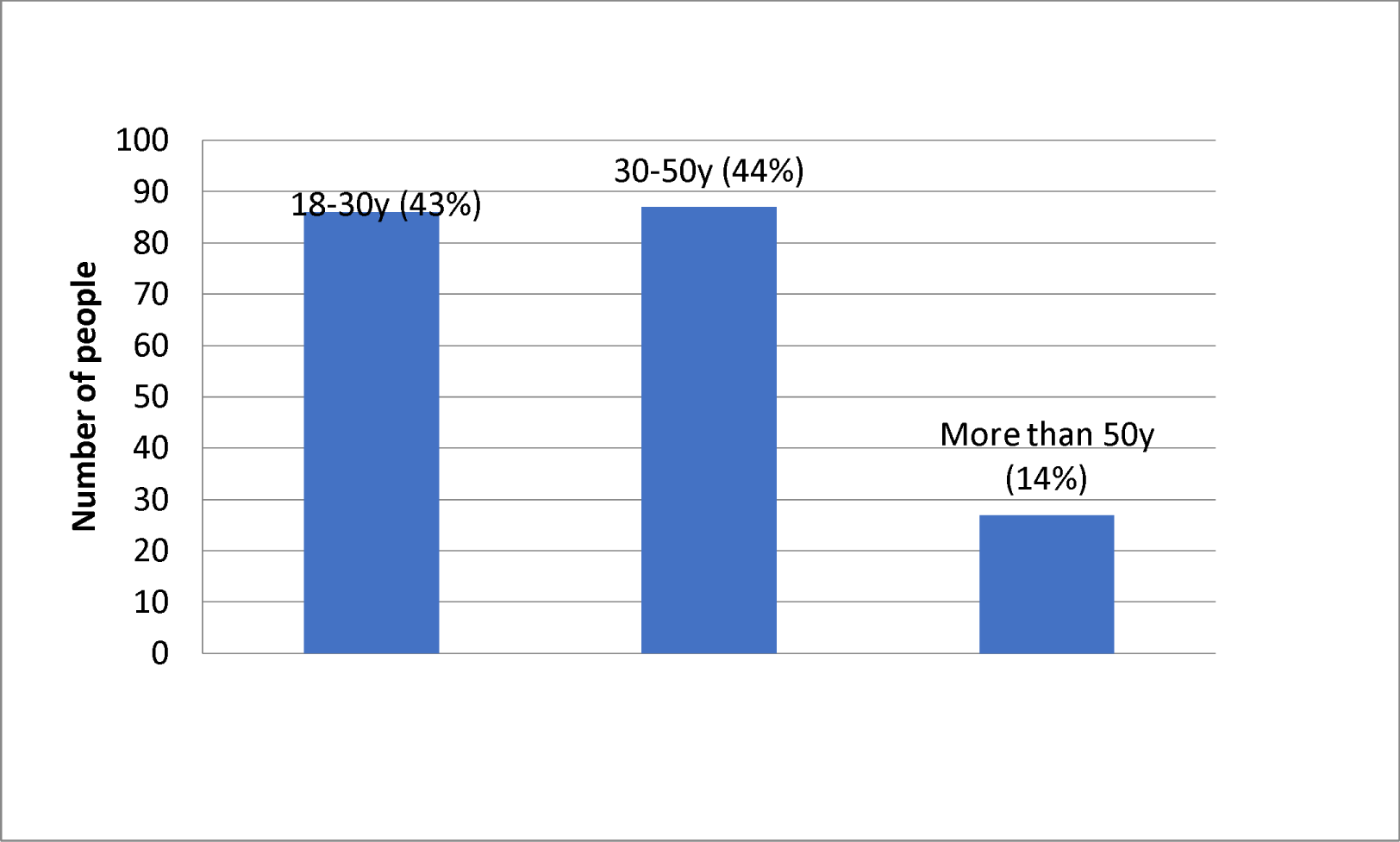
Age

Regarding the level of education, it is seen that of the total of 200 people interviewed, only 4% (n=8) have completed tertiary education; 25% (n=50) completed secondary education; 55% (n=110) only finished primary school and 16% (n=32) did not finish any type of study. In this graph (Graph 3) you can see how the population has a tendency to only complete primary studies.

**Graf. 3.**
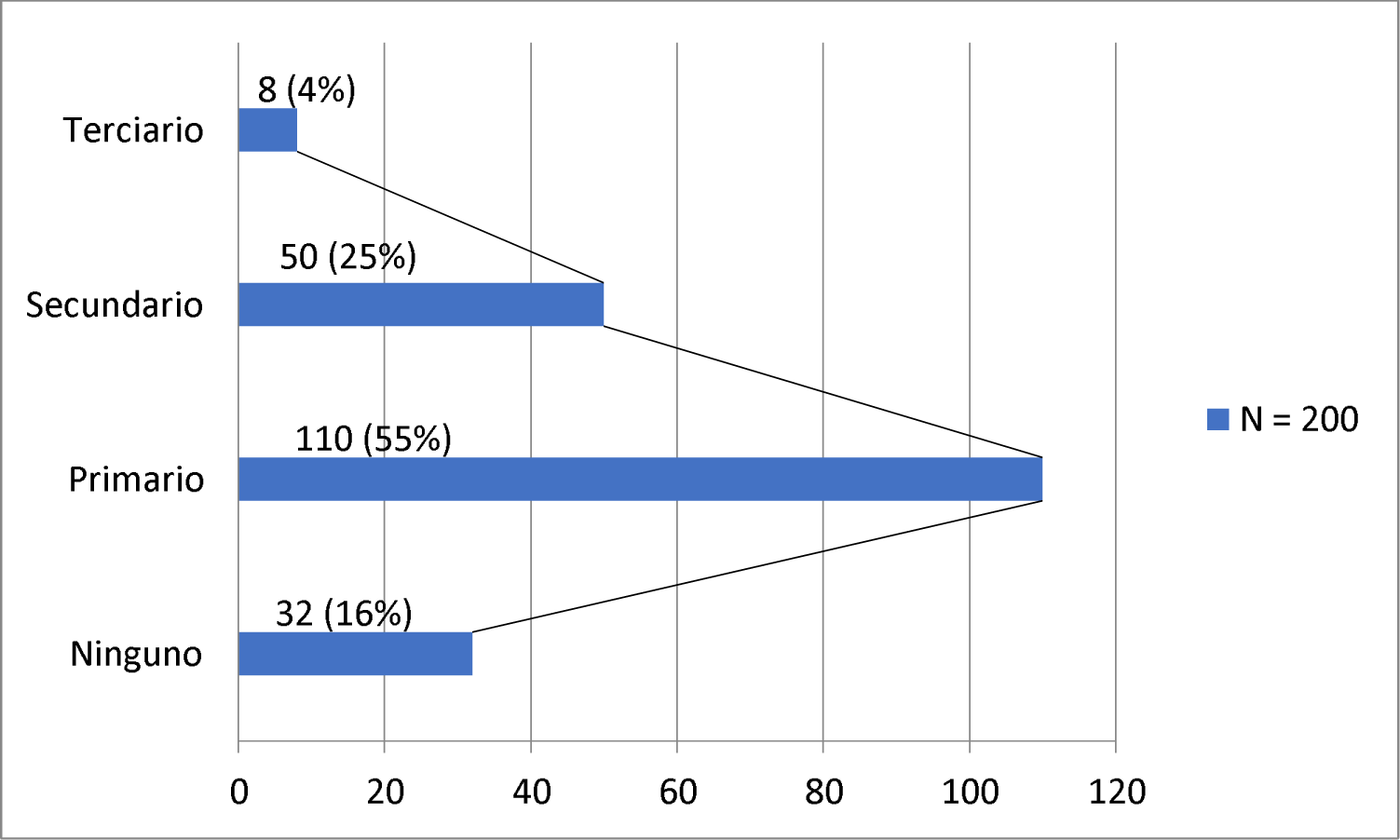
Educational level

Of the total patients, 12% (n=23) live with more than 8 people; 49% (n=97) live with between 5 and 8 people, and 40% (n=80) between 1 and 4 people. The average number of people they live with is 6 people (Graph 4).

**Graf. 4.**
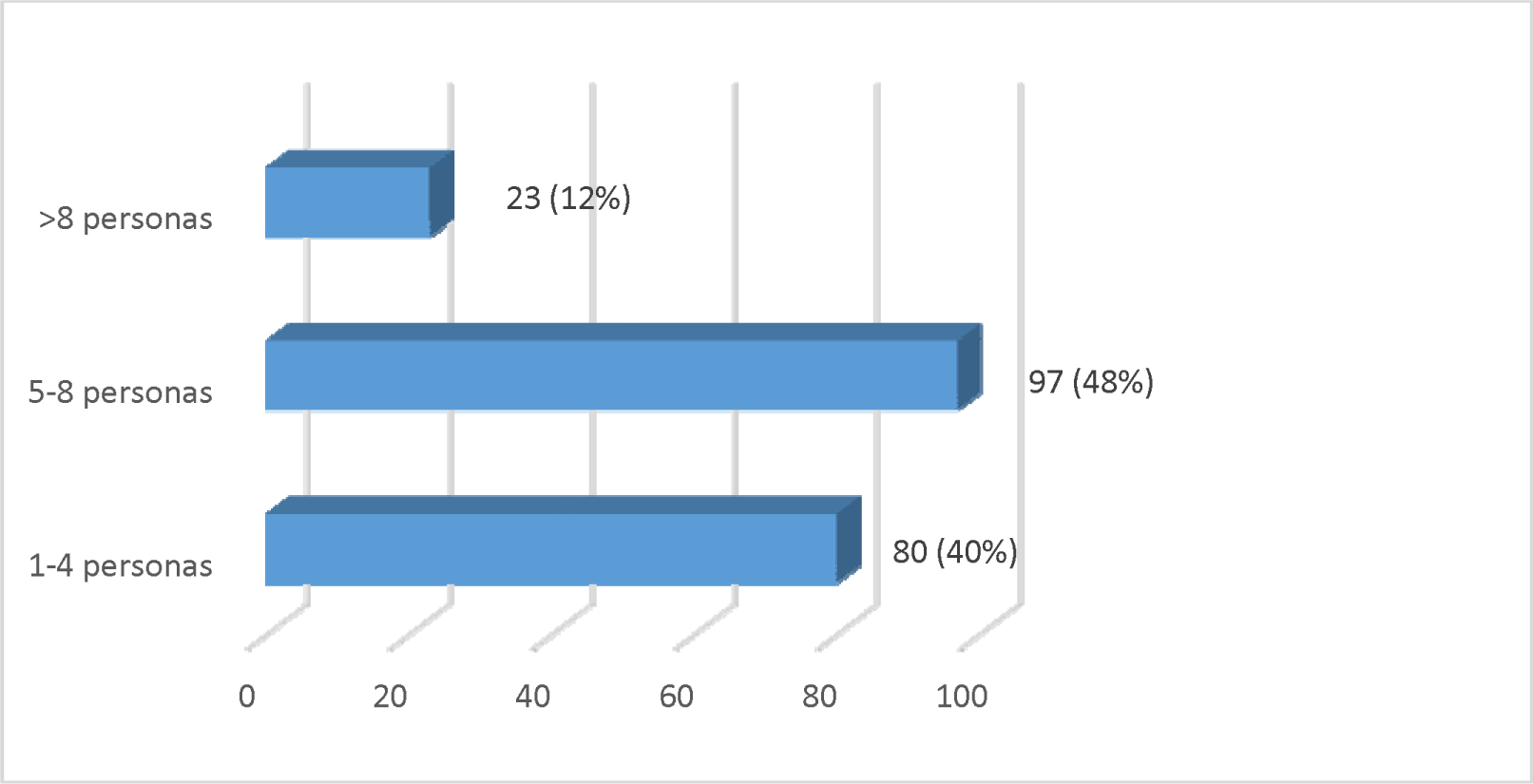
People per Household

Of the total number of people interviewed, their occupations were: unemployed/do not answer 18% (n=36); laborer 4% (n=7); cleaning 4% (n=7); worker/bricklayer 6% (n=11); municipal employee 6% (n=12); changas 6% (n=11); student 5% (n=9) and housewife 45% (n=89).

**Graf. 5.**
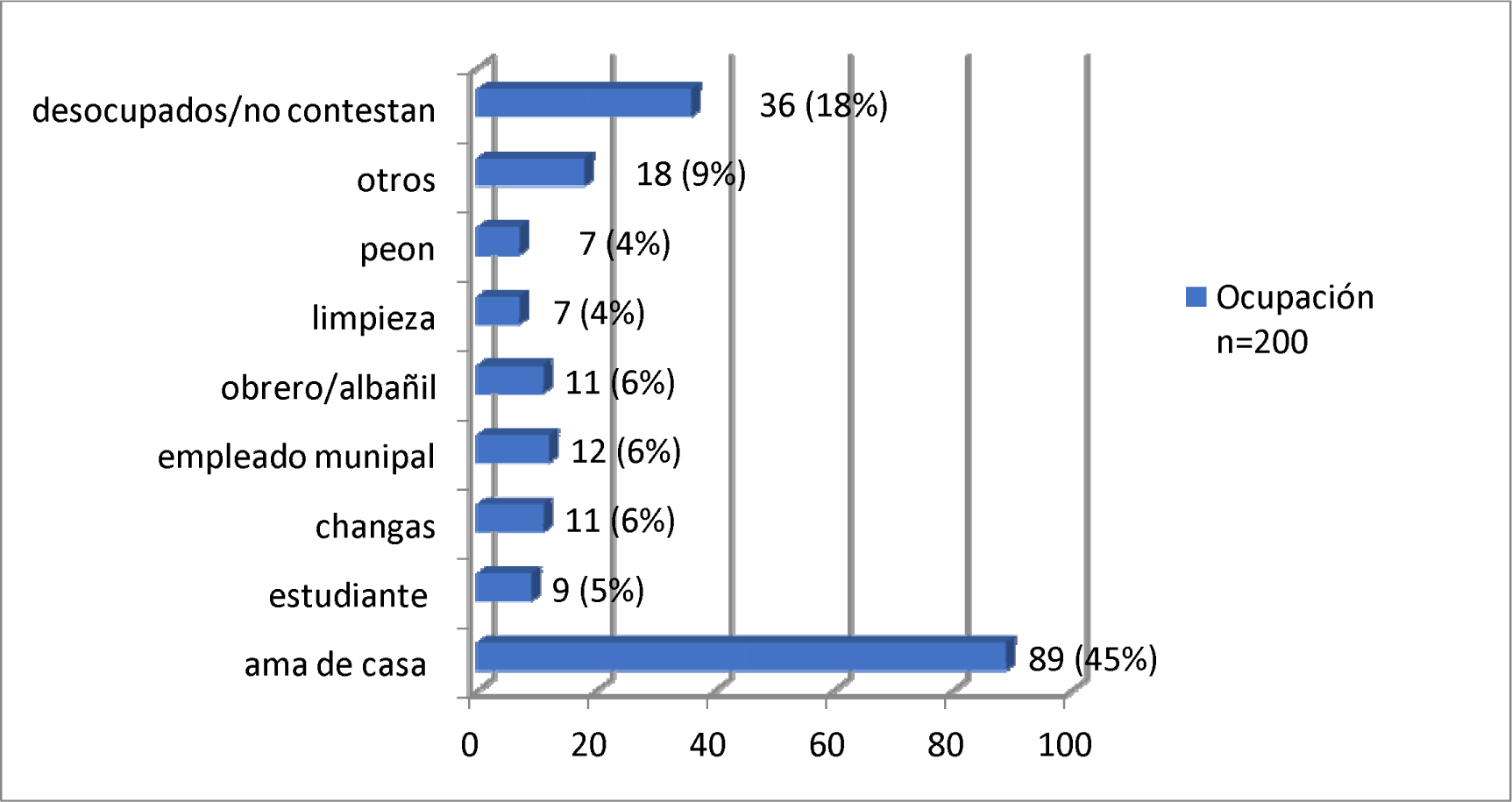
Occupations

Among those interviewed, 12% (n=24) presented social work; 1% (n=1) had a private plan; 12% (n=9) had a public plan and 79% (n=158) did not have any of these.

**Graf. 6.**
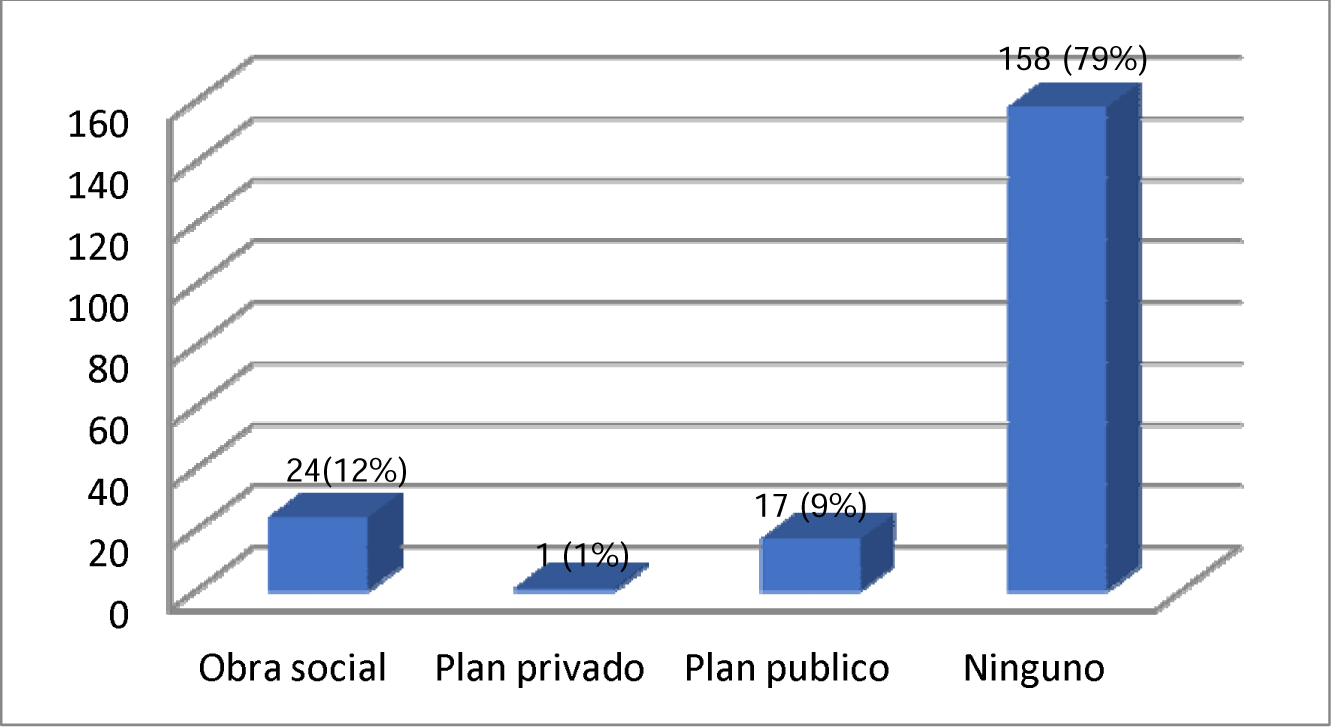
Health Coverage

In the following graph **Graph 7** you can see the places of residence of the people interviewed. Only the neighborhoods with a sample of more than 5 respondents are diagrammed below, resulting in: Neighborhood 120 homes 3% (n=5); Colonia Osvaldo 7% (n=11); Stingy 8% (n=13); the Triangle 4% (n=6); Manzione 8% (n=12); the Leñera 8% (n=13); the Falklands 11% (n=17); Worker 5% (n=8); Dora Colony 5% (n=7); Canal Melero 4% (n=7); pink field 8% (n=12); Place 3% (n=5).

**Graf 7.**
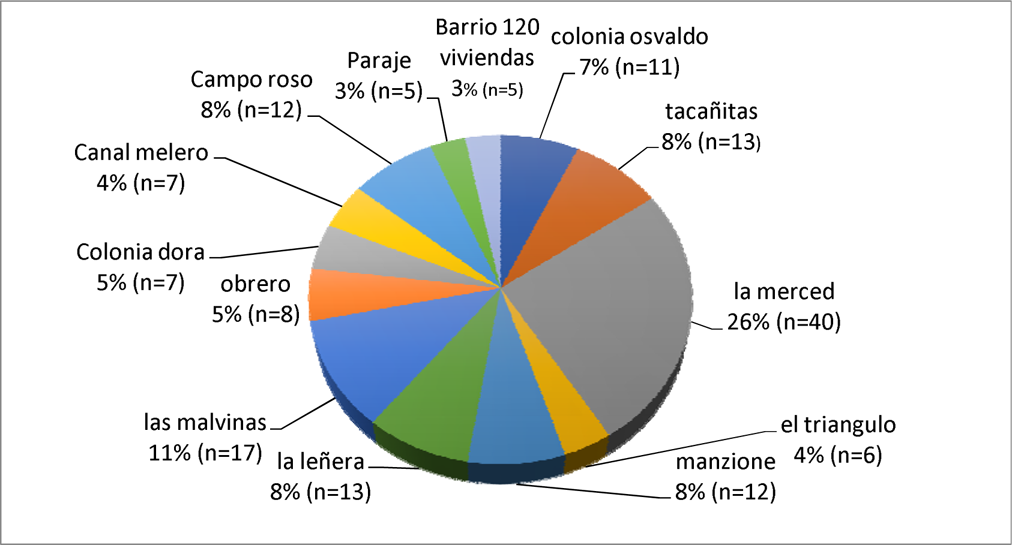
Address. Only the neighborhoods where more than 5 interviewees lived are shown.

Below (Graph 8) you can see how of the total number of respondents (n=200), 70% (n=61) answered NO to the question of whether they live with a person with Chagas disease, while 30% (n=61) I assure you YES.

**Graf. 8.**
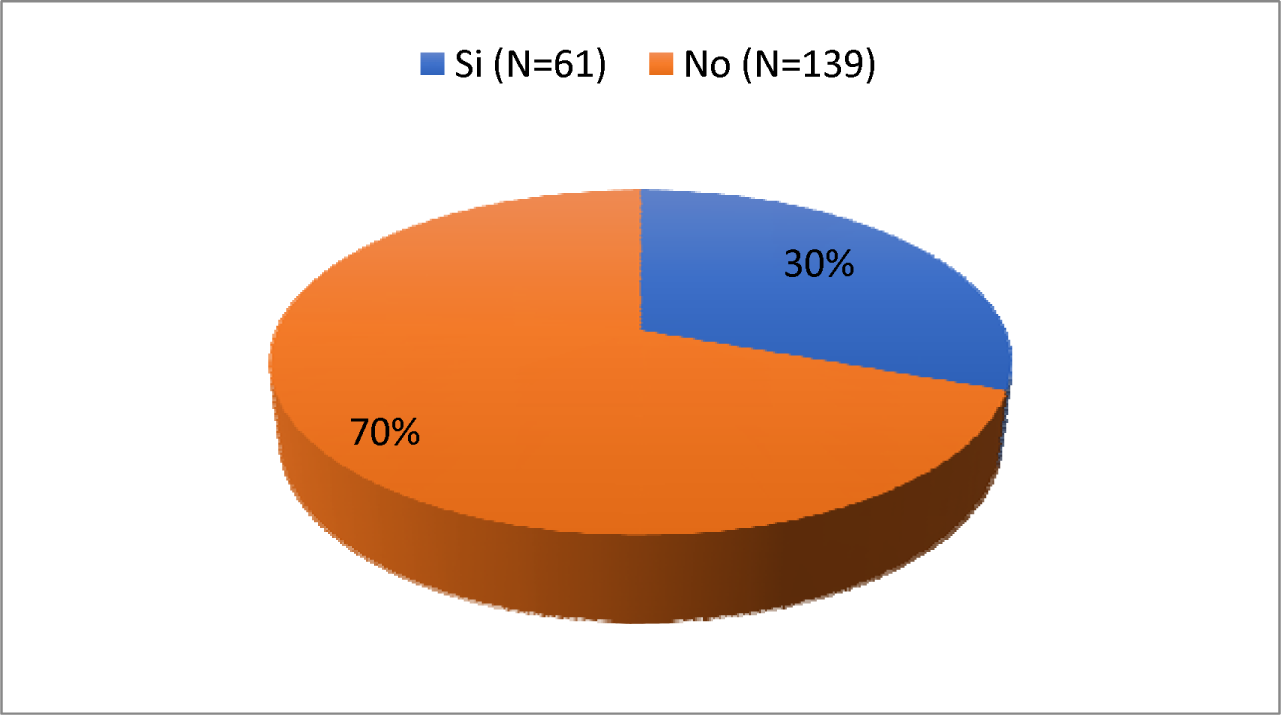
Do you live with anyone with Chagas?

In this graph (Graph 9) you can see how of those interviewed (n=200) 84% (n=167) had pets while only 16% (n=33) answered no to this question. Graph 9.1 shows that of the 167 people who had pets, 58% (n=161) had dogs; 20% (n=56) cats; 15% (n=43) chickens; others (goats, horses, pigs, cows) 7% (n=19). Finally, graph 9.2 shows the number of pets per household, with 34% having only one pet and 63% having more than two pets per household.

**Graf. 9.**
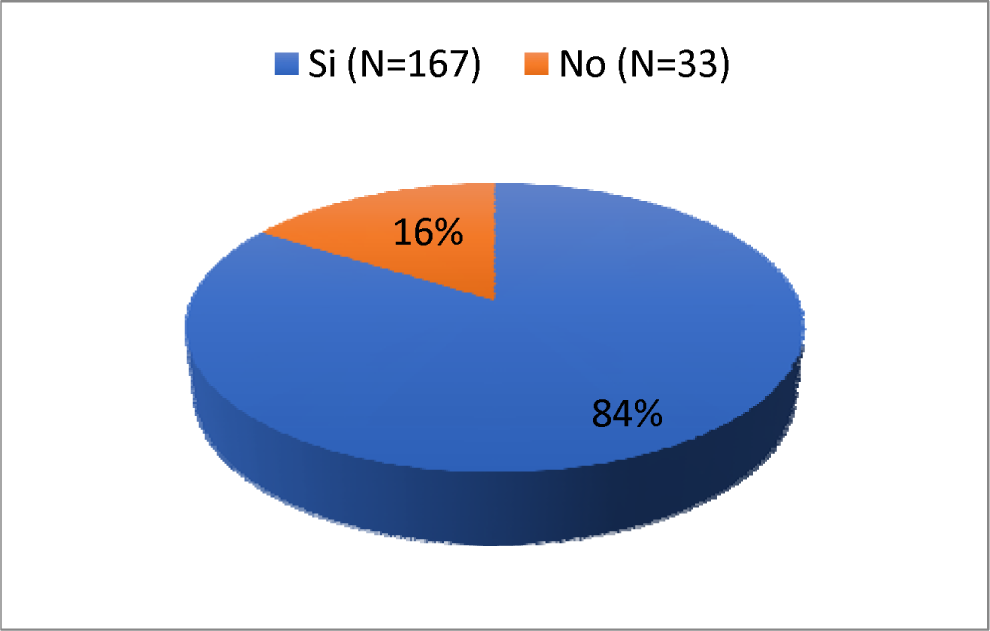
Do you have pets?

**Graf. 9.1.**
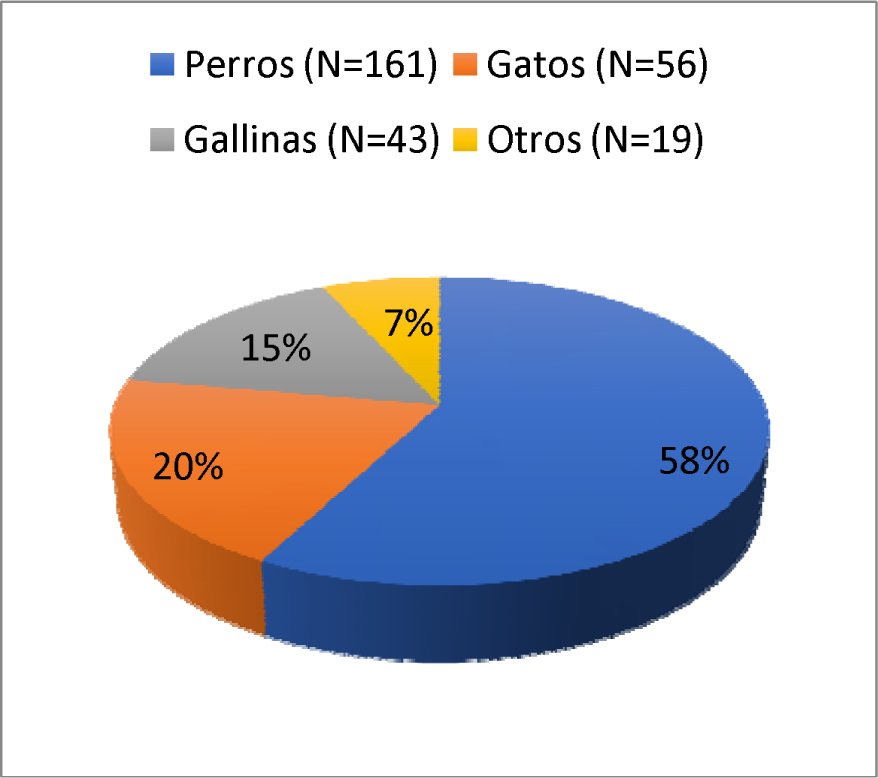
Types of Pets

**Chart 9.2.**
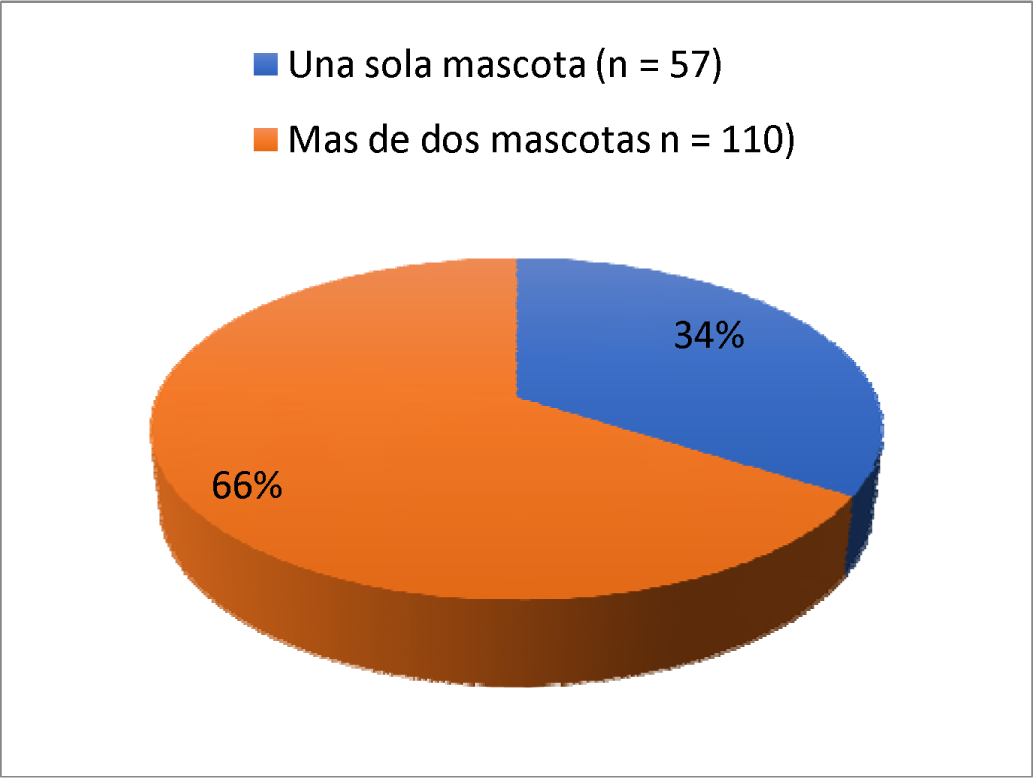
Number of pets per household

The following graph (Graph 10) summarizes whether the housing was made of material, to which of the 200 interviewed, 22% responded NO while 78% responded YES. Of the total, 84% answered YES to the question of whether they had electricity in their home, while 16% answered NO. 68% responded that they DID have drinking water, while 32% responded that they did not have it. Regarding waste collection, 49% of those surveyed responded that YES they collected it, while 51% responded NO.

**Graf 10.**
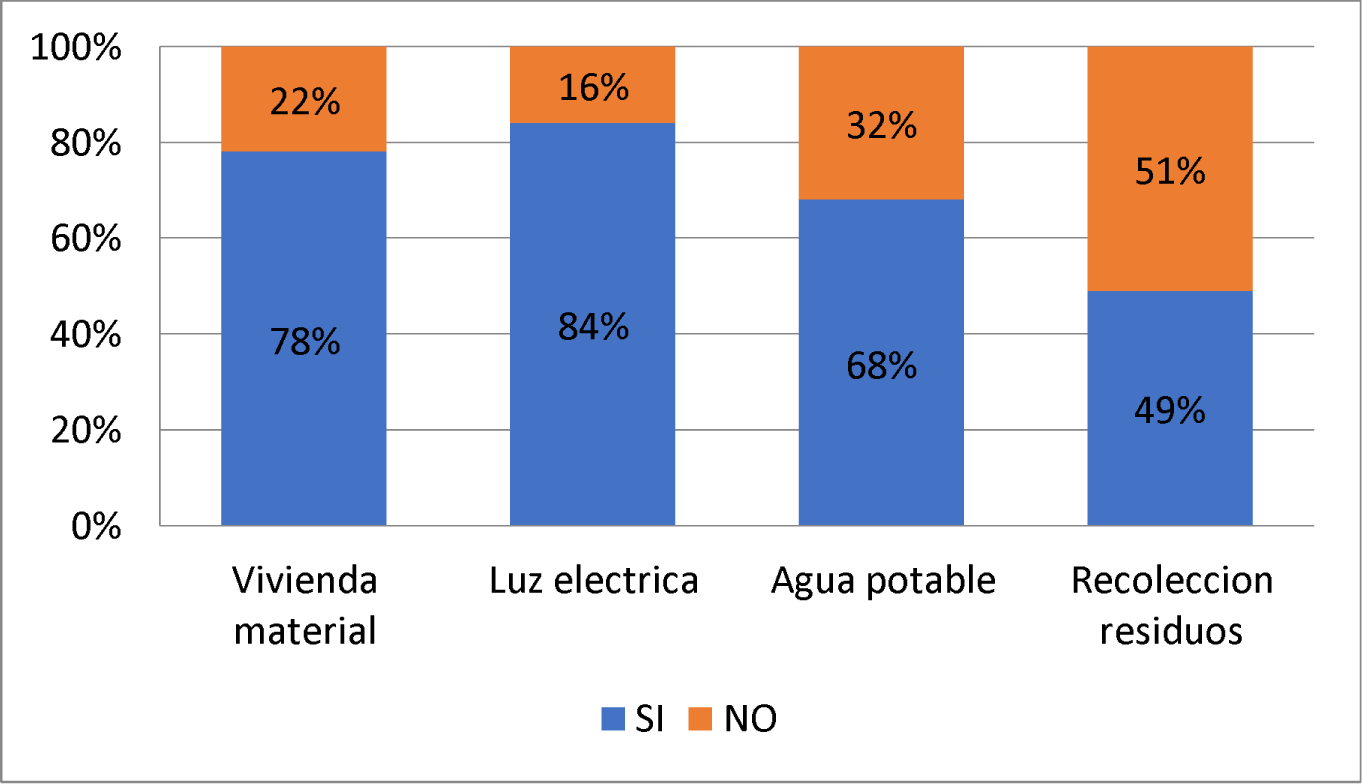
Housing

Below (Graph 11) it can be seen that of the total number of respondents (n=200), the difference between those who have ever witnessed a vinchuca or traces in their home with those who have not is minimal, stating that 51% DID witness them (n = 87) and NO 49% (n = 113). However, 45% (n=91) claim to have ever witnessed vinchucas or traces of it in their home, while the remaining 55% (n=109) did not (Graph 12).

**Graf 11.**
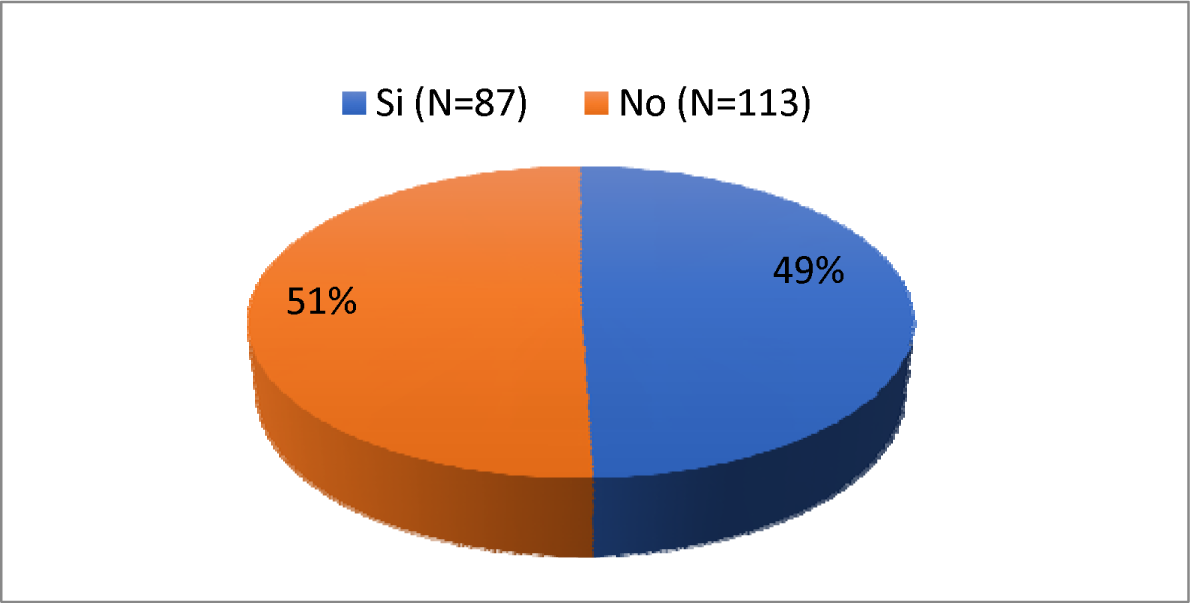
Have you ever witnessed a vinchuca or traces (feces, molts or eggs) of a vinchuca in your home?

**Graf 12.**
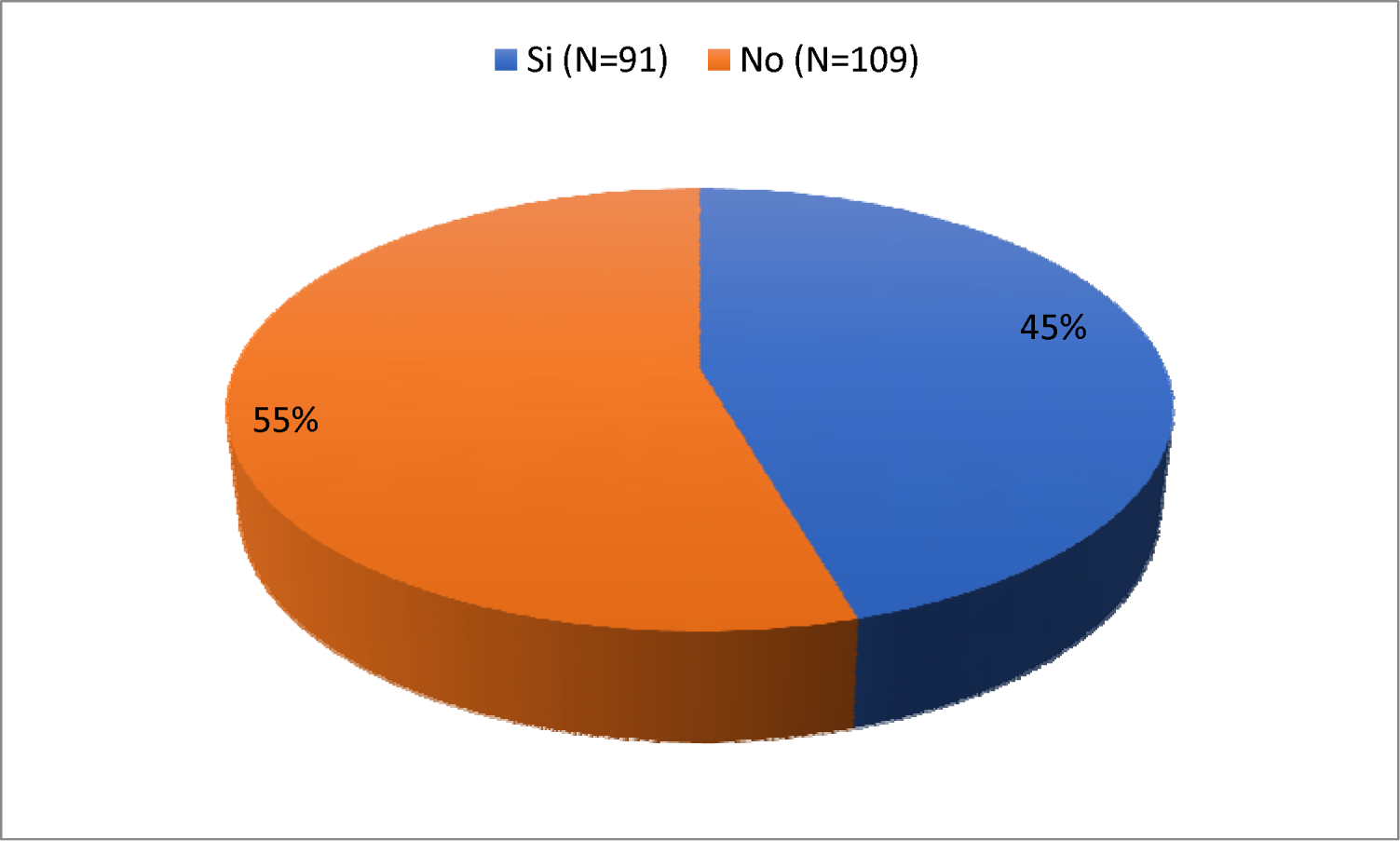
Have you ever witnessed a vinchuca or traces of a vinchuca around your home?

In this graph (Graph 13) about fumigation in their home or surroundings, it can be seen that of the total number of respondents (n=200), 77% (n=154) answered YES to the question while only 23% (n =46) answered NO.

**Graf. 13.**
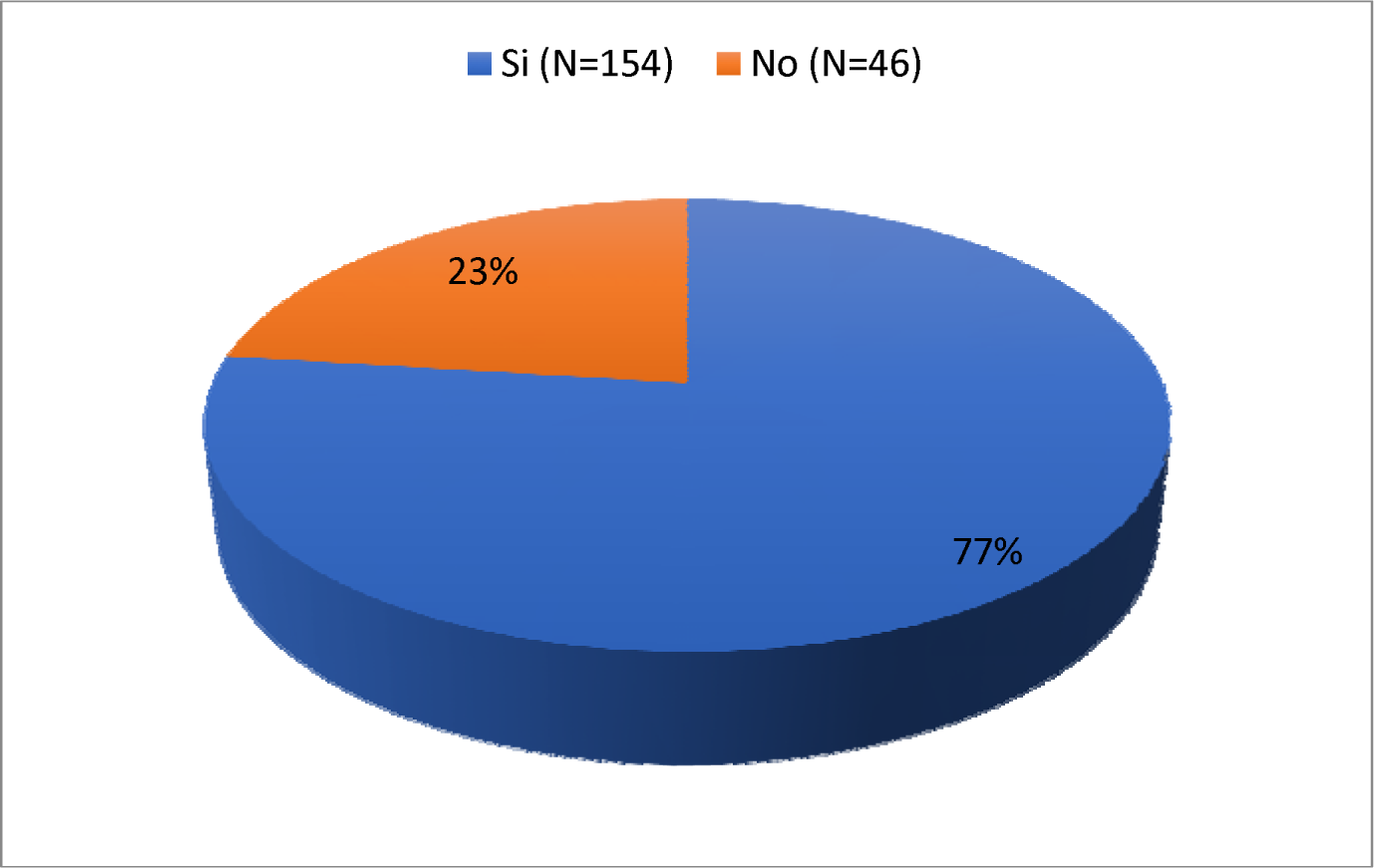
Have you ever fumigated your home or surrounding areas?

In the following graph (Graph 14) about the frequency of fumigation, it can be seen that of the total number of people who received fumigation at home (n=154), 62% (n=61) responded that they had it done once a year. year, that is, annually; 19% every 6 months, that is, semi-annually; 9% every 1-6 months and 10% in a period greater than 1 year.

**Graf. 14.**
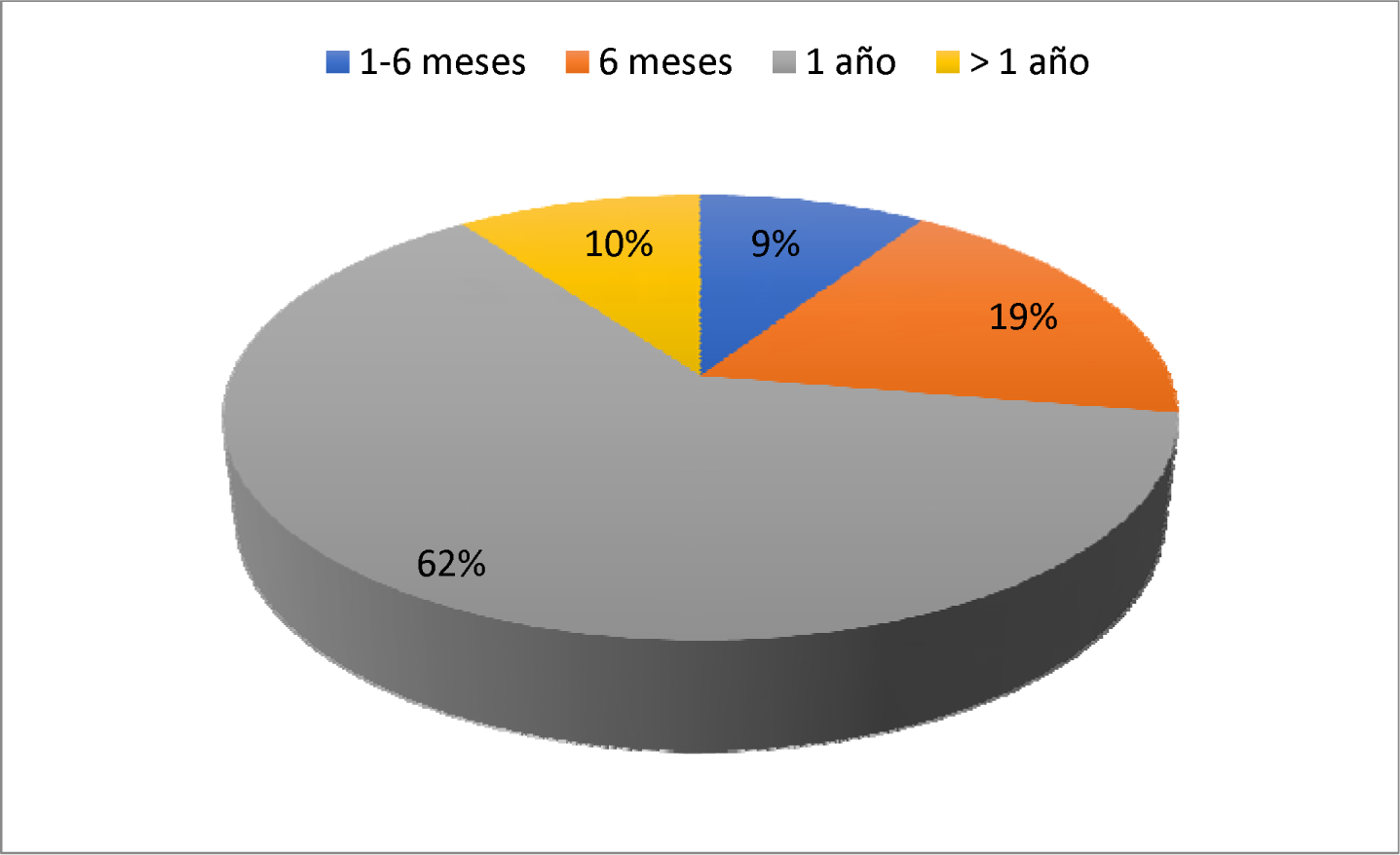
Fumigation frequency

In relation to the patients who had risky housing (n = 43), that is, ranch type (thatched/mud/canvas/sheet metal roof, mud/cracked walls), we can see that the highest percentage has risky roofs and walls. 98% (n = 42) and 77% (n = 33) respectively, as well as 90.6% (n = 39) have chicken coops, dogs or birds in their home or peridomicile.

**Graf 15.**
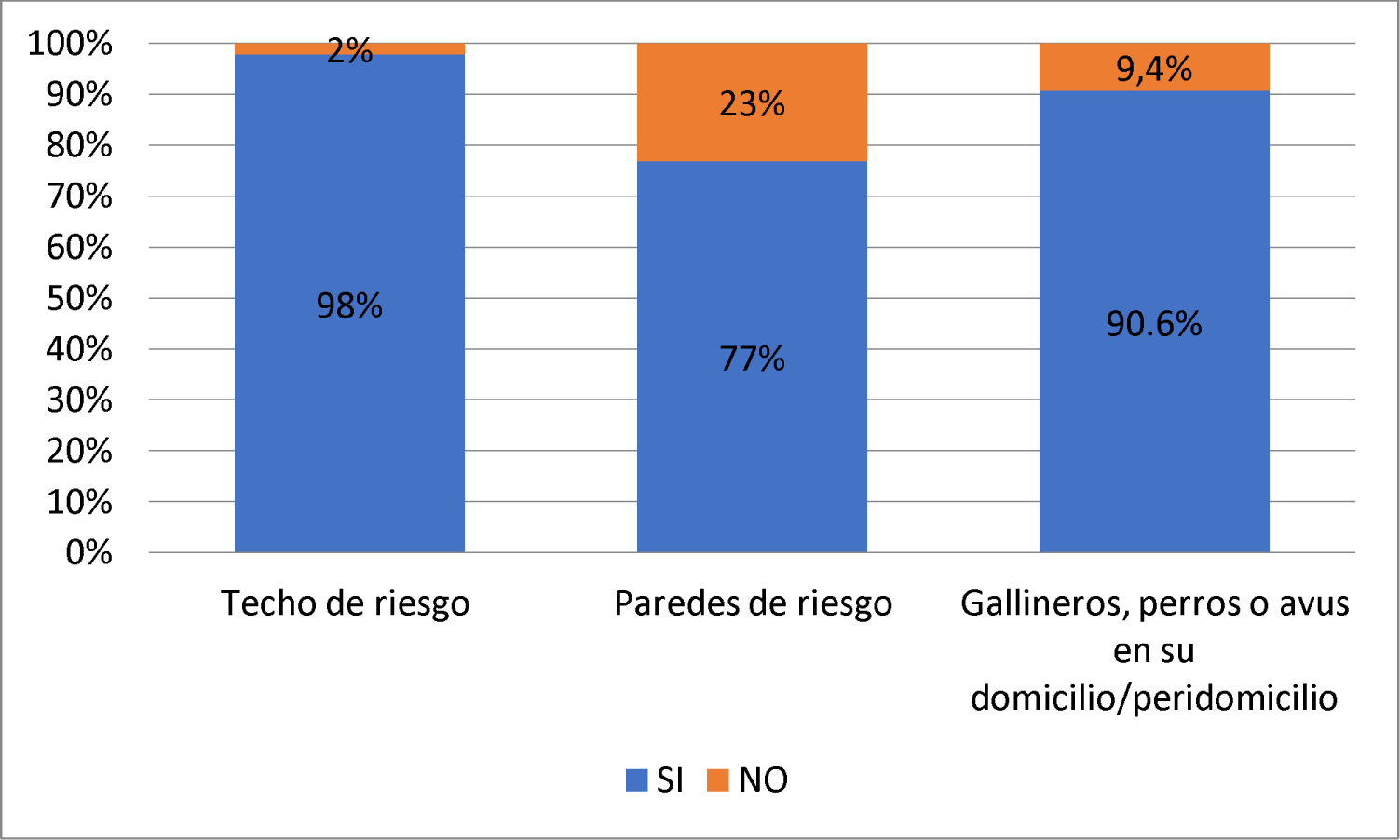
Risk housing.

Results of the second section surveyed: BASIC KNOWLEDGE ABOUT CHAGAS DISEASE

In the following graph (Graph 16) you can see how of those interviewed (n=200) 32% (n=64) did not recognize adult vinchucas while 68% (n=136) did recognize them.

**Graf 16.**
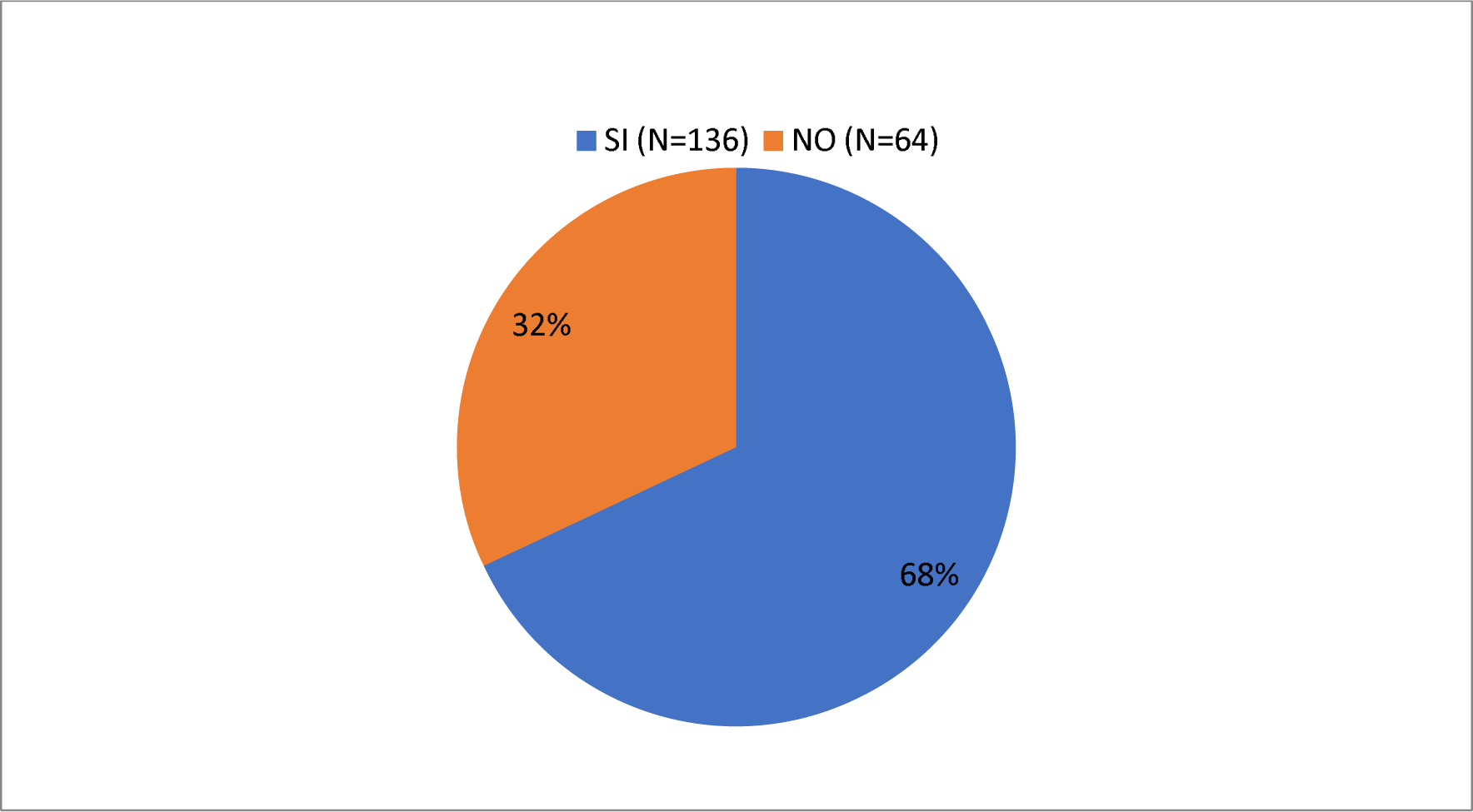
Do you recognize adult vinchucas?

In this graph (Graph 17) you can see how of those interviewed (n=200) 59% (n=118) did not recognize the presence of vinchuca feces on the walls while 41% (n=82) responded yes. they recognized them

**Graph 17.**
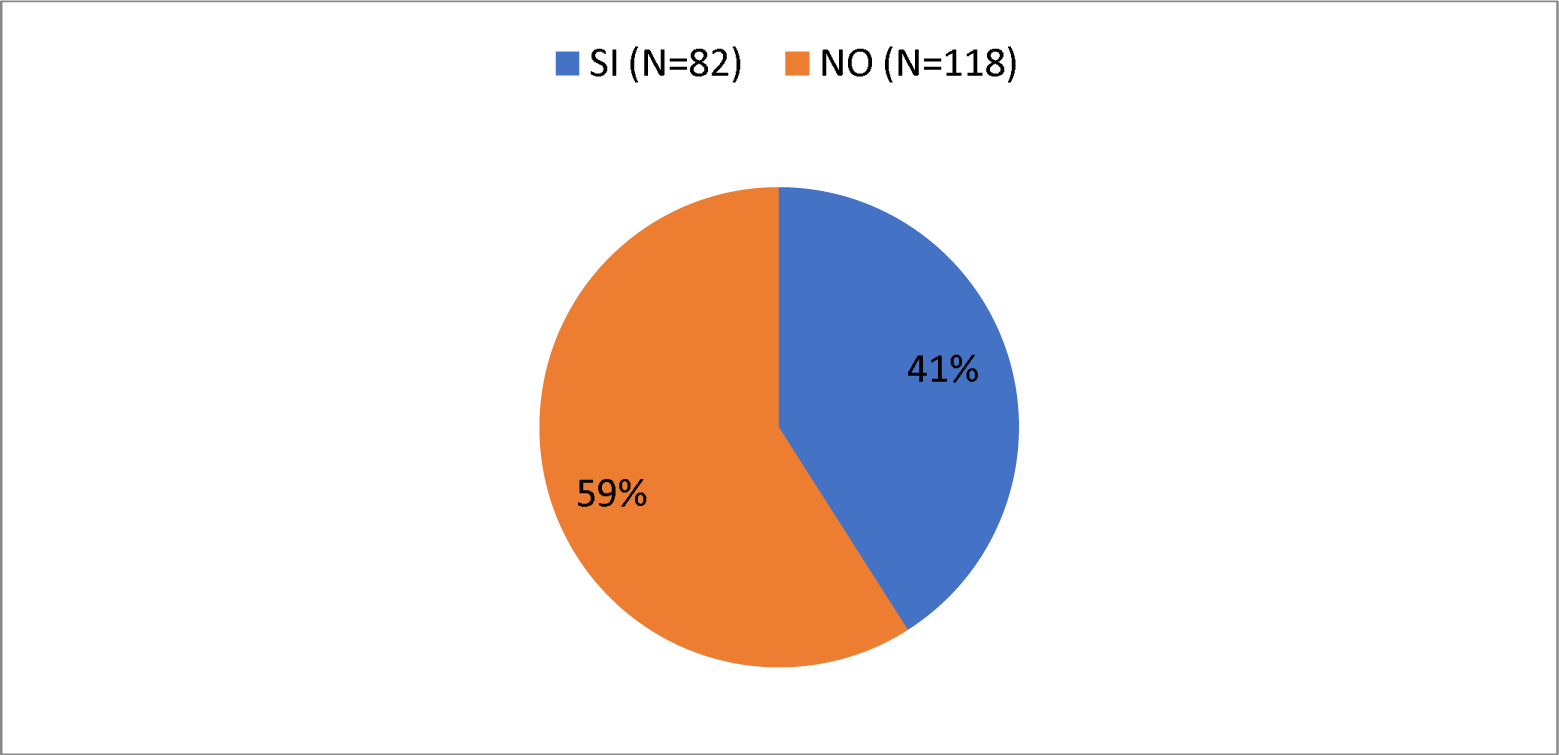
Do you recognize the presence of vinchucas feces on the walls?

To continue, in this graph (Graph 18) you can see how of those interviewed (n=200) 56% (n=112) answered Yes to this question, while 44% (n=88) answered No. to the same.

**Graf 18.**
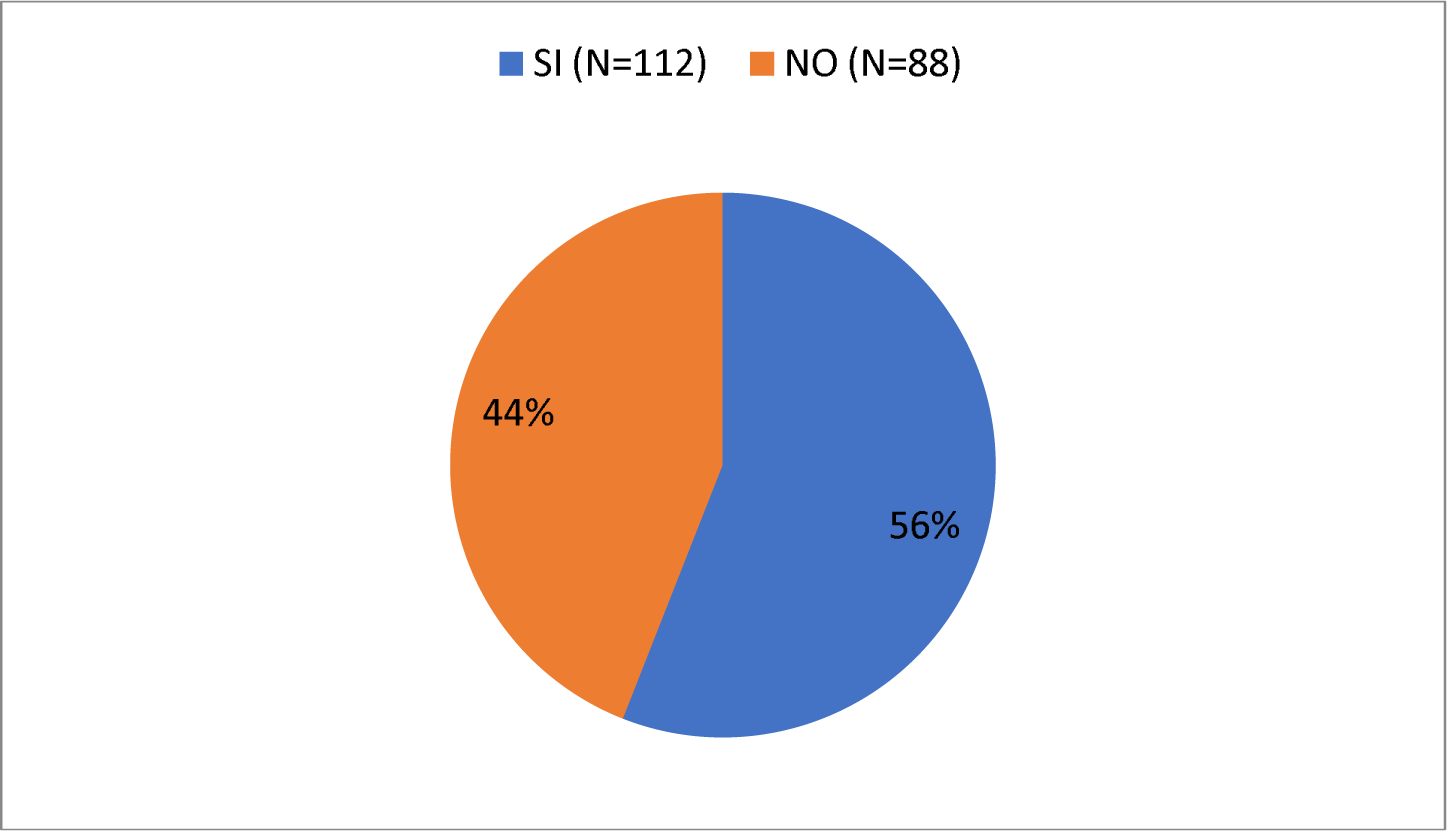
Do you know that the most common habitat for vinchucas in your home is the kitchen and the bedroom?

In this following graph (Graph 19) you can see how of those interviewed (n=200) 80% (n=160) responded that if they were aware that the vinchuca can be housed in chicken coops, pens and material warehouses, while 20% (n=40) responded that they were not aware.

**Graf 19.**
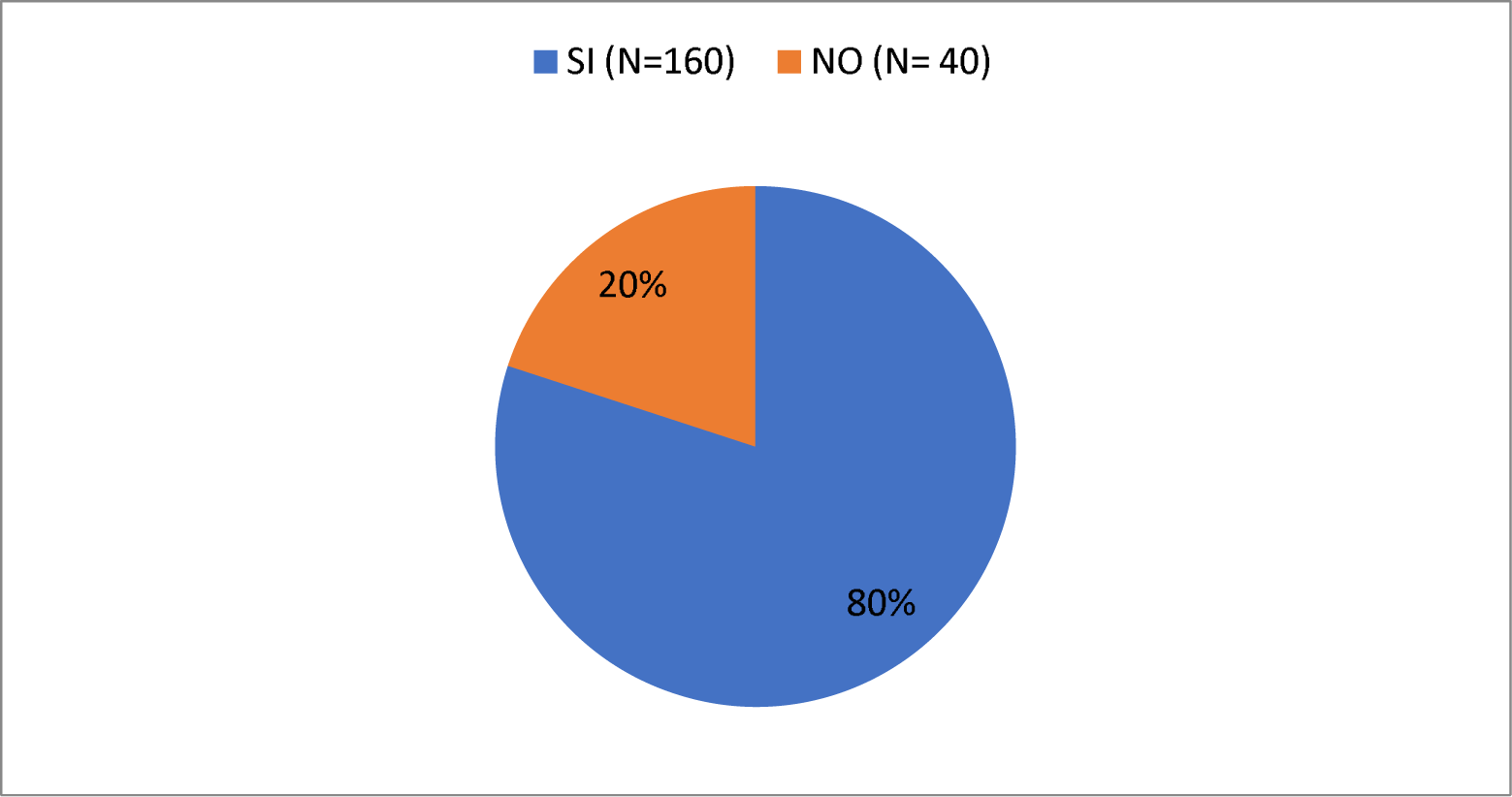
Do you know that outside your home, the vinchuca can stay in chicken coops, pens and material storage?

In relation to whether they had knowledge about the vinchuca shelters, it can be seen that of those interviewed (n=200) 80% (n=161) answered Yes to this question while 20% (n=39) responded than No to it (Graph 20).

**Graf. 20.**
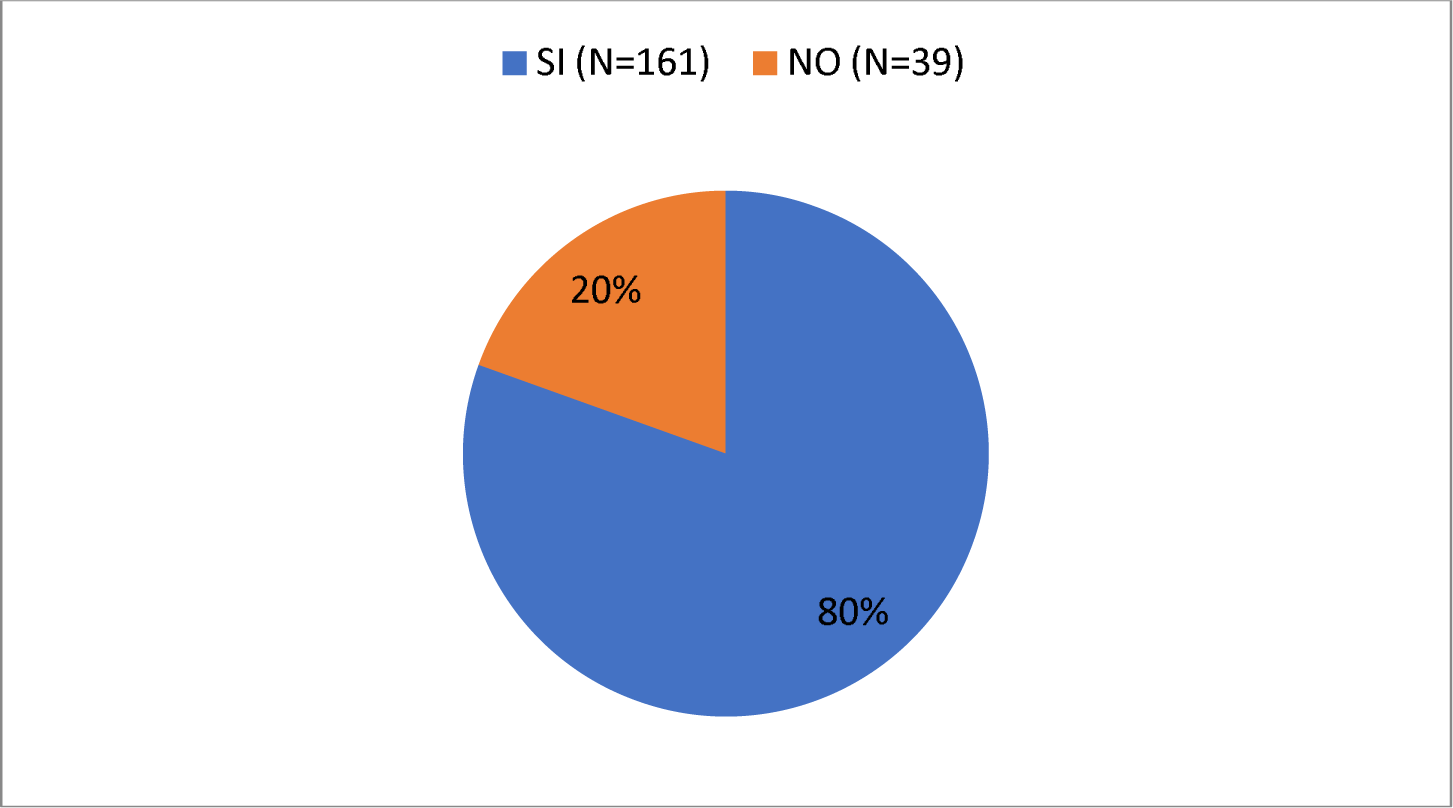
Do you know that the vinchuca’s refuges are in the wall, ceiling, under the bed and in cracks?

Below (Graph 21) you can see how of those interviewed (n=200) 72% (n=145) responded that YES they were aware that disorder favors the presence of vinchucas, while 28% (n=55) They responded NO to it.

**Graf 21.**
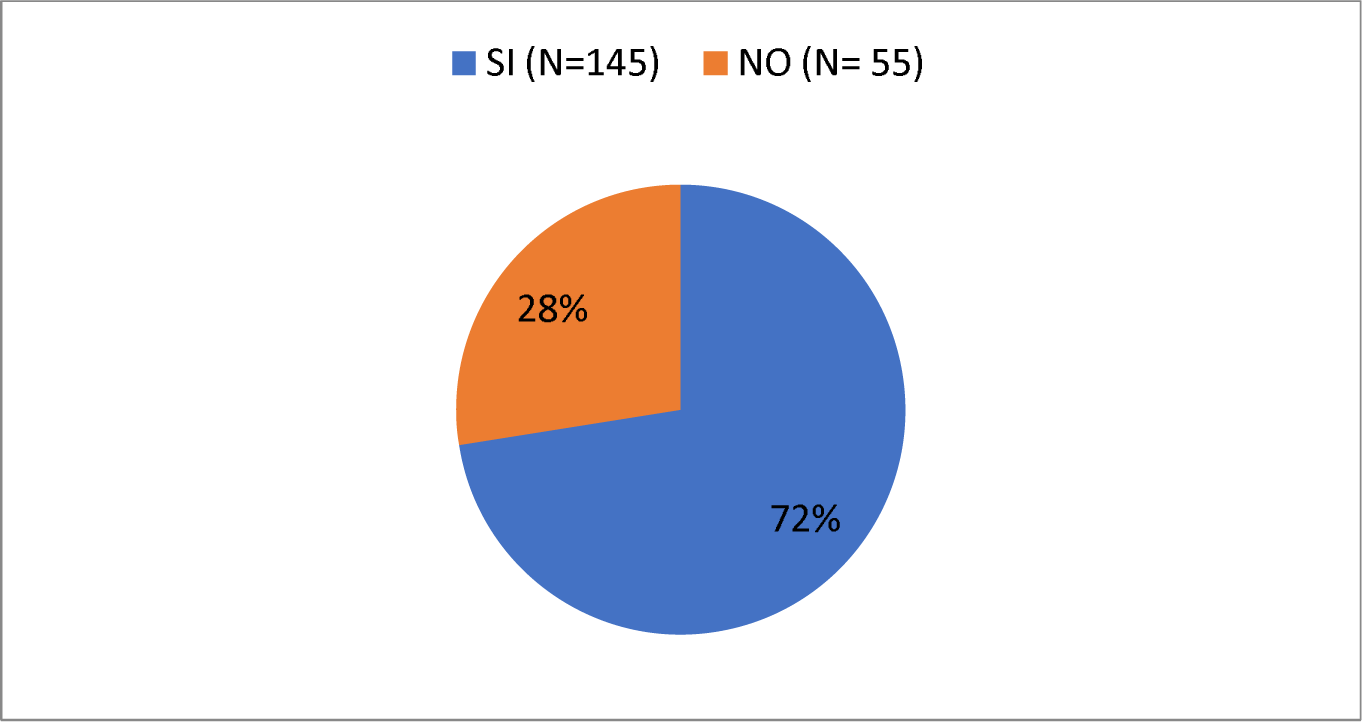
Do you know that disorder favors the presence of vinchucas?

In this graph (Graph 22) it can be seen how of those interviewed (n=200) 95% (n=190) responded that they DID know that ranch-type housing favors the presence of vinchucas, while 5% (n =10) responded NO to it.

**Graf 22.**
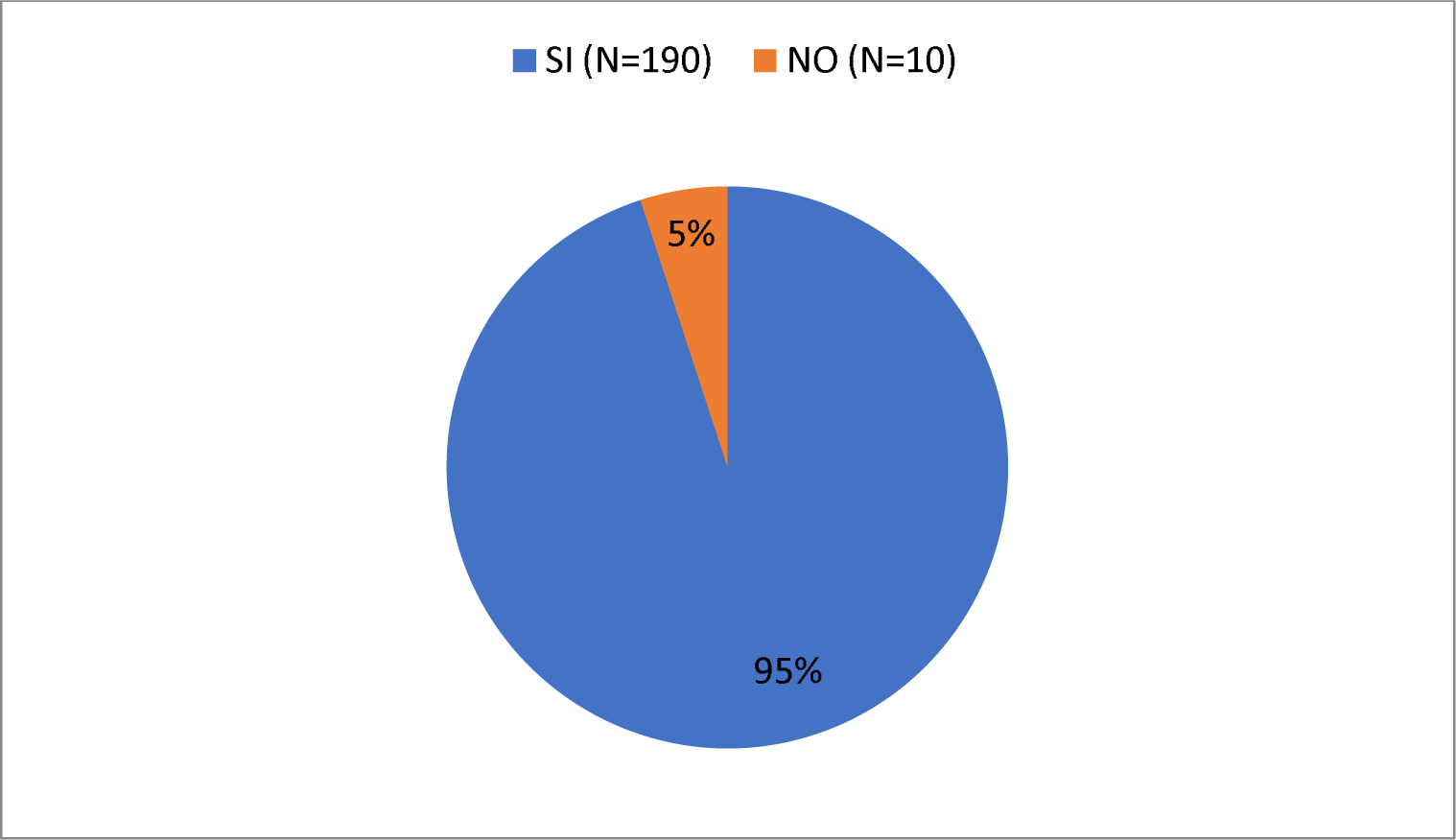
Do you know that ranch-type homes favor the presence of vinchucas?

In this graph (Graph 23) you can see how of those interviewed (n=200) 62% (n=124) answered Yes to this question while 38% (n=76) answered No to it.

**Graf 23.**
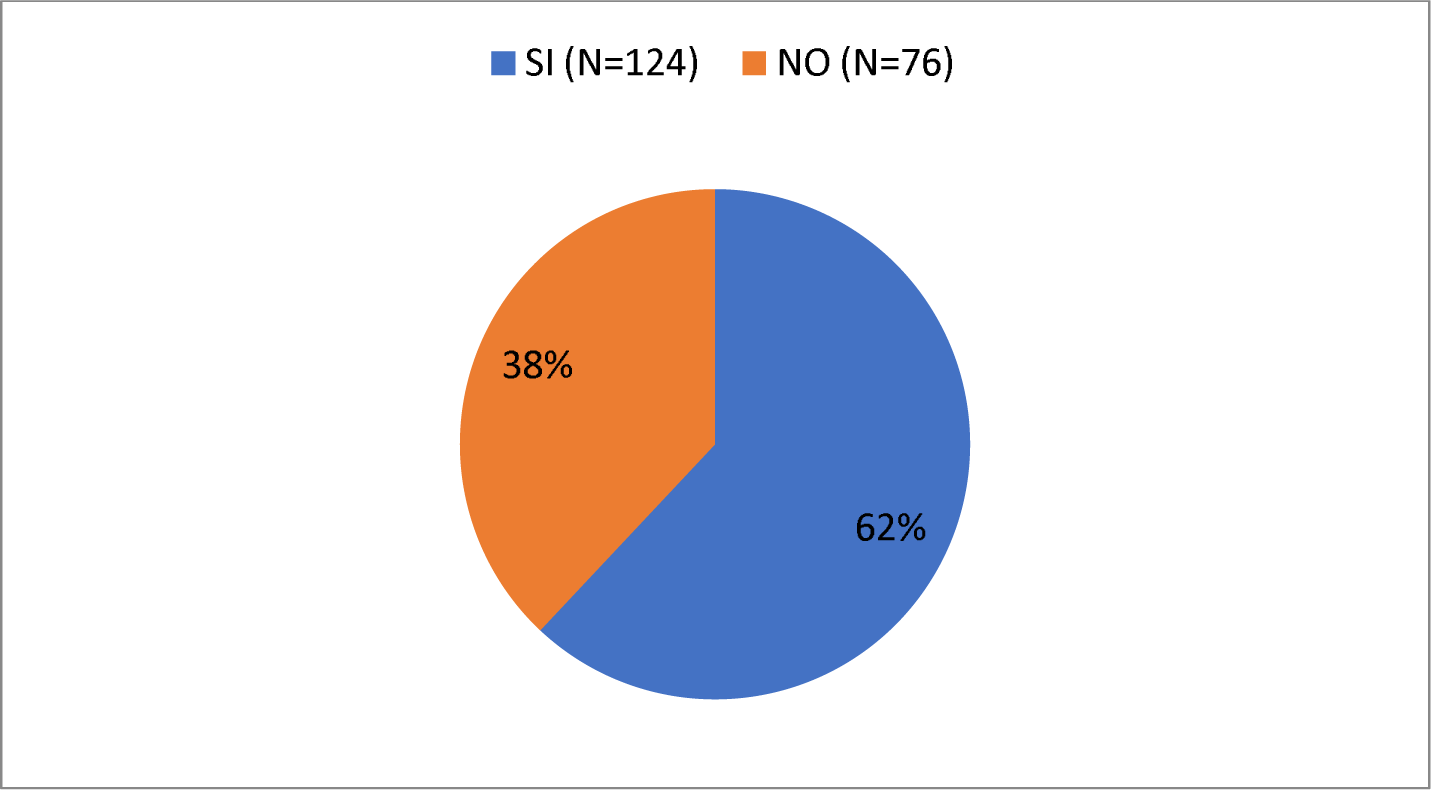
Do you know that the number of vinchucas increases in summer?

In this graph (Graph 24) you can see how of those interviewed (n=200) 91% (n=182) answered Yes to this question while 9% (n=18) answered No to it.

**Graf 24.**
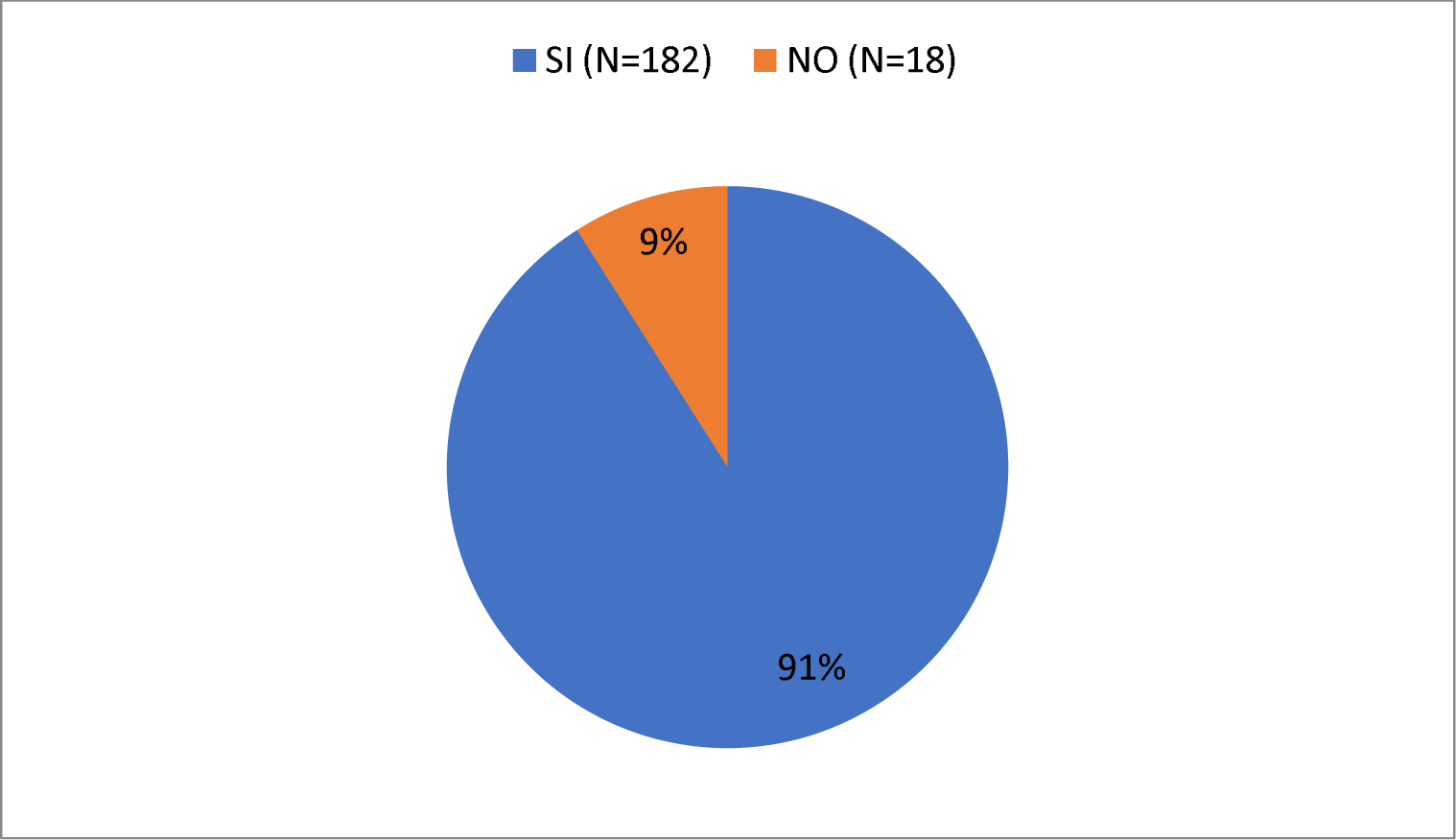
Do you know that the vinchuca feeds on blood?

Regarding the question: “Do you know that vinchucas bite at night?” It can be seen that of those interviewed (n=200) 79% (n=159) responded that Yes they were aware and 21% (n=41) responded that No. (Graph 25)

**Graph 25.**
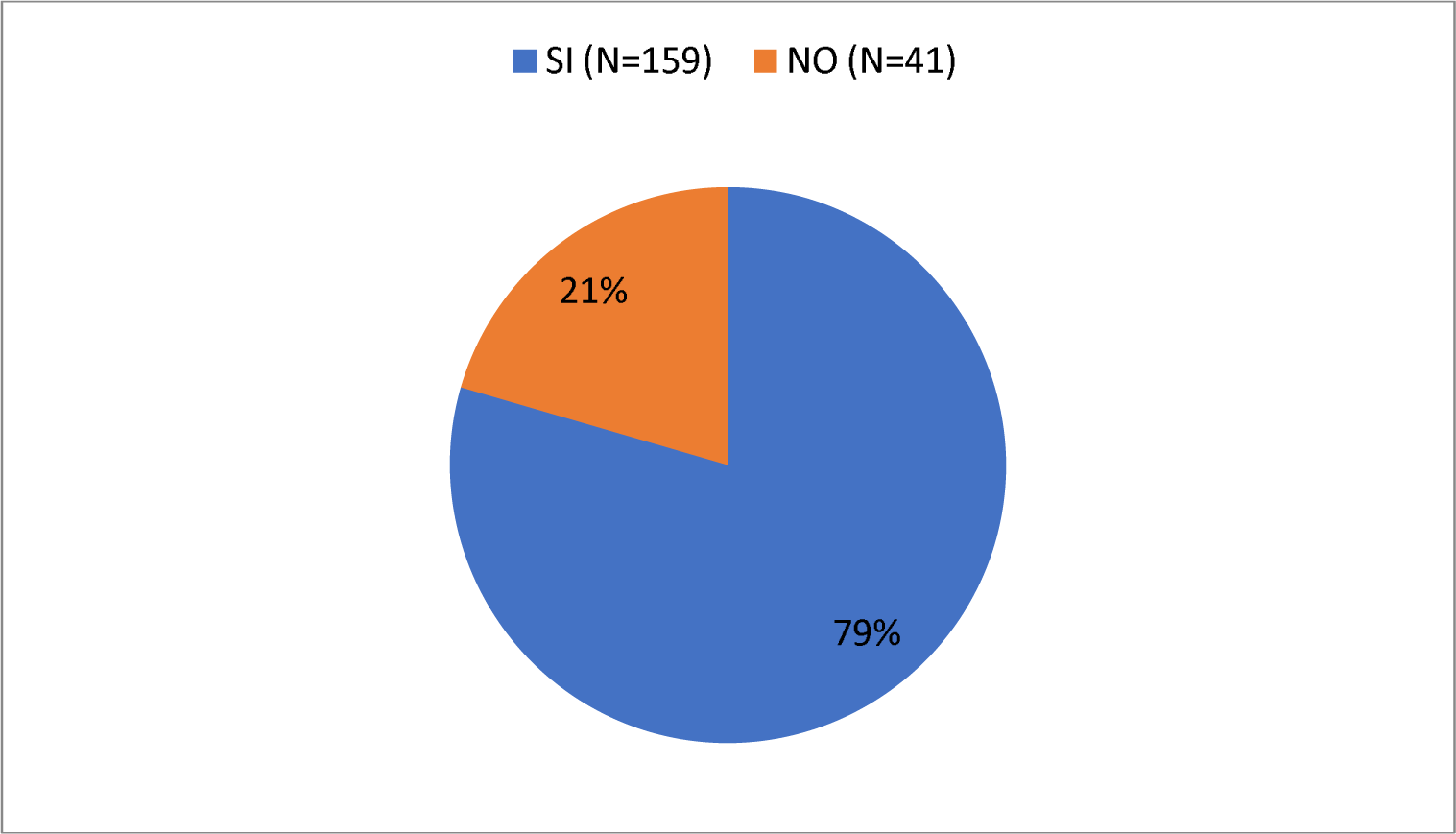
Do you know that vinchucas bite at night?

In this graph (Graph 26) you can see how of those interviewed (n=200) 96% (n=192) answered Yes to this question while 4% (n=8) answered No to it.

En este gráfico *(Gráfico 26)* se puede observar como de los entrevistados (n=200) un 96% (n=192) respondieron que Si a esta pregunta mientras que un 4% (n=8) respondieron que No a la misma.

**Graph 26.**
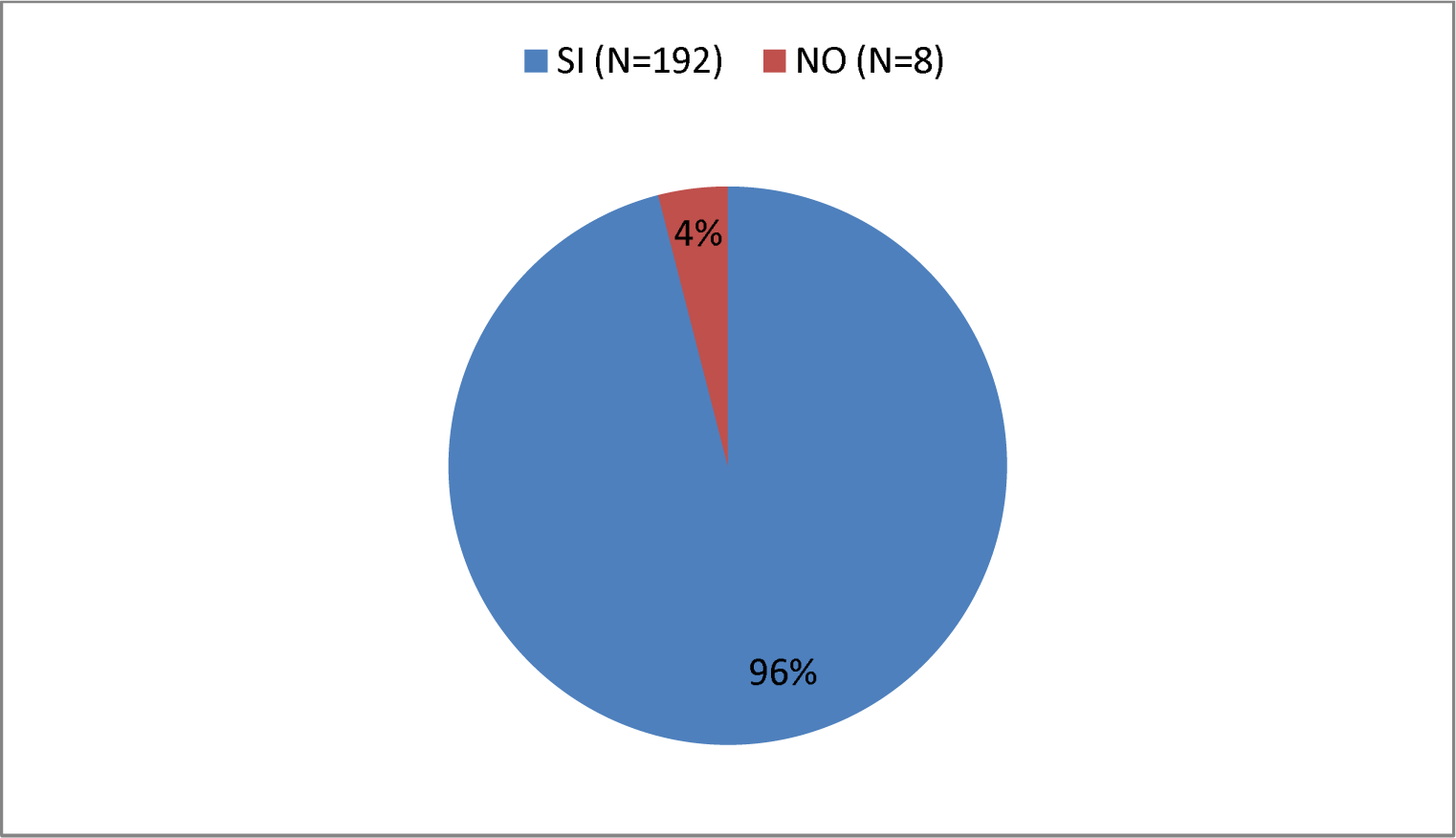
Do you know that vinchucas bite humans?

In this graph (Graph 27) you can see how of those interviewed (n=200) 70% (n=141) answered Yes to this question while 30% (n=59) answered No to it.

**Graph 27.**
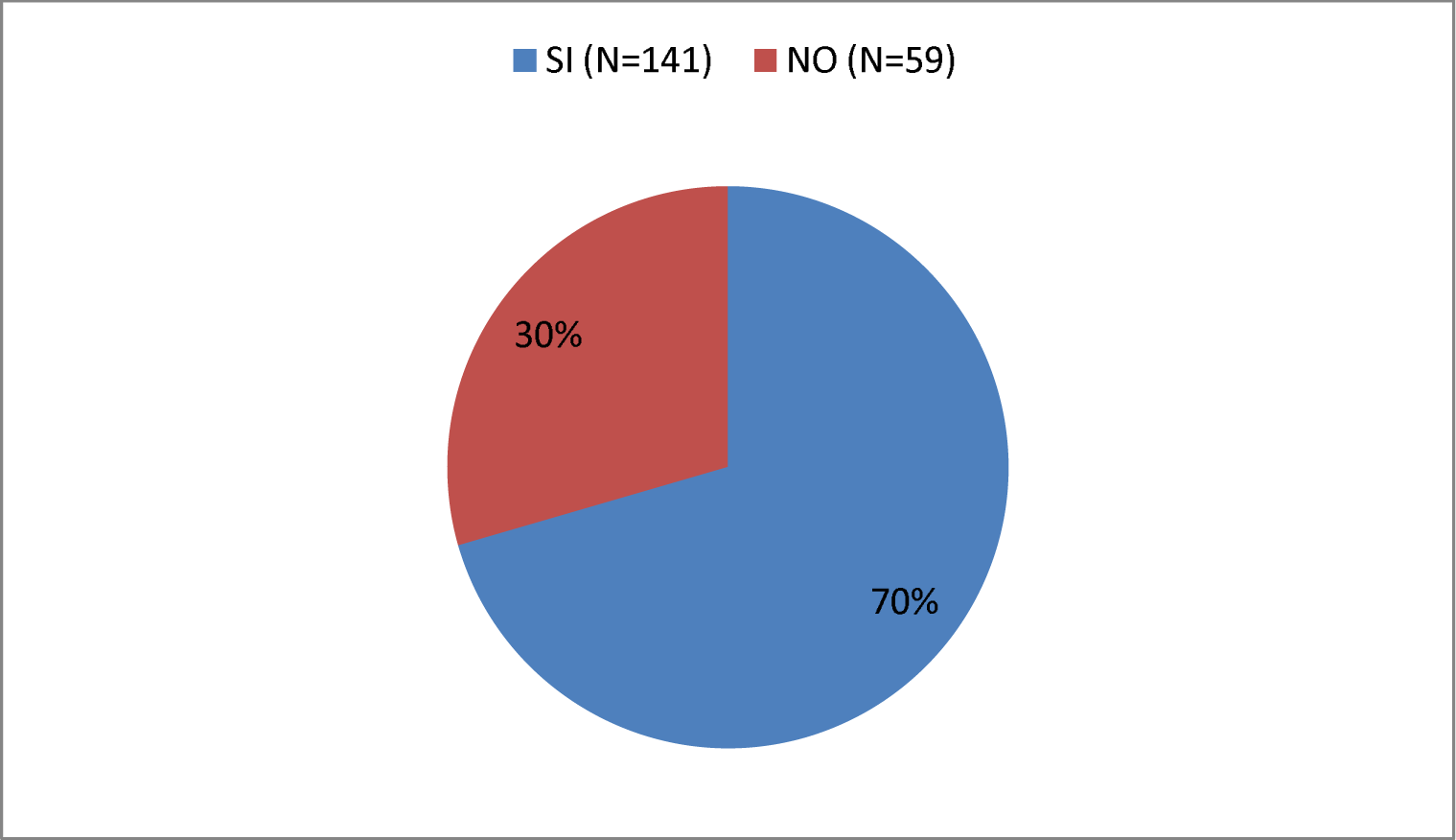
Do you know that vinchucas bite chickens, birds and other mammals?

Below (Graph 28) you can see how of those interviewed (n=200) 89% (n=179) answered Yes to this question while 11% (n=21) answered No to it.

**Graph 28.**
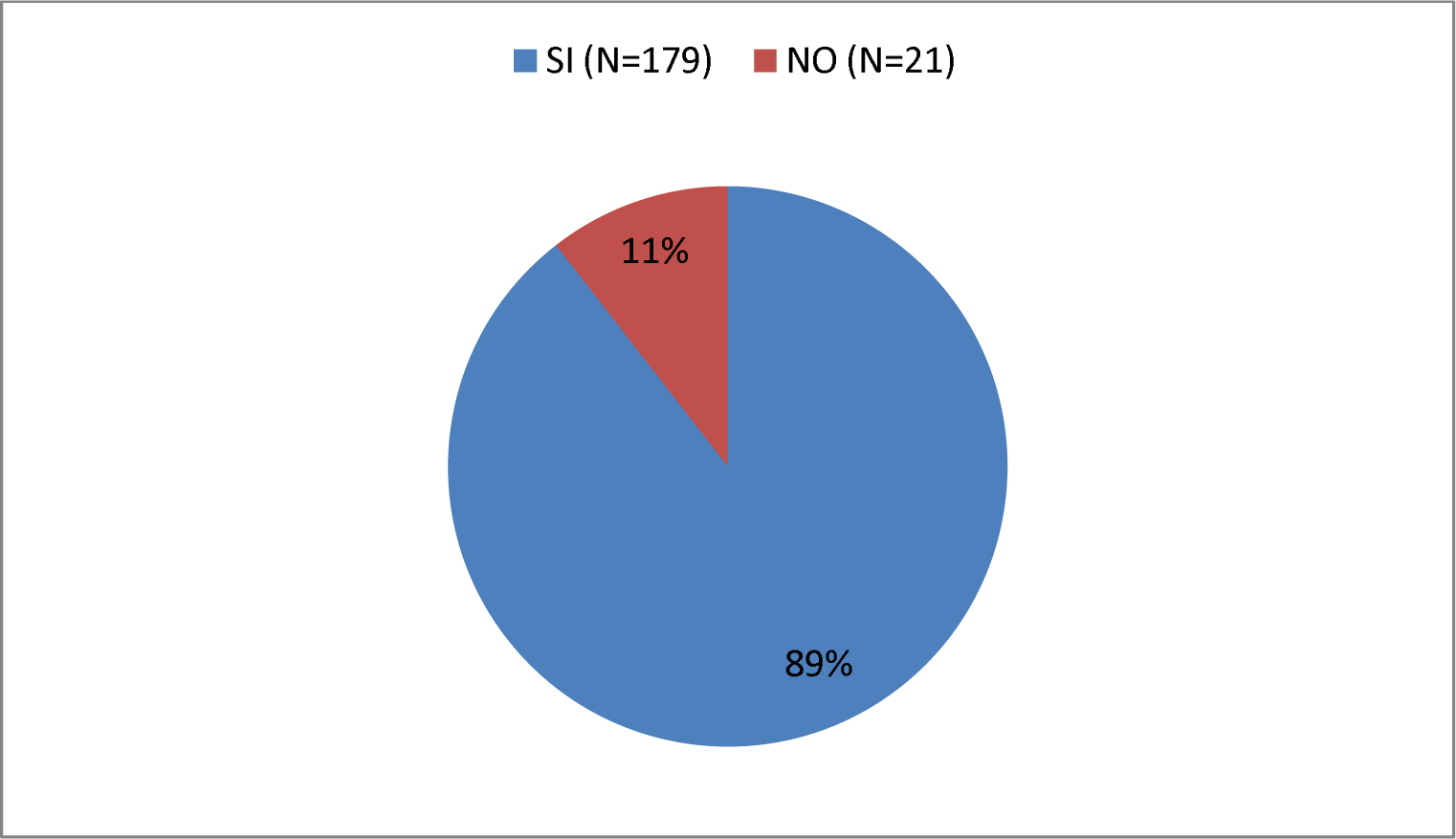
Do you know that vinchucas transmit a disease?

In this graph (Graph 29) you can see how of those interviewed (n=200) 80% (n=160) answered Yes to this question while 20% (n=40) answered No to it.

**Graph 29.**
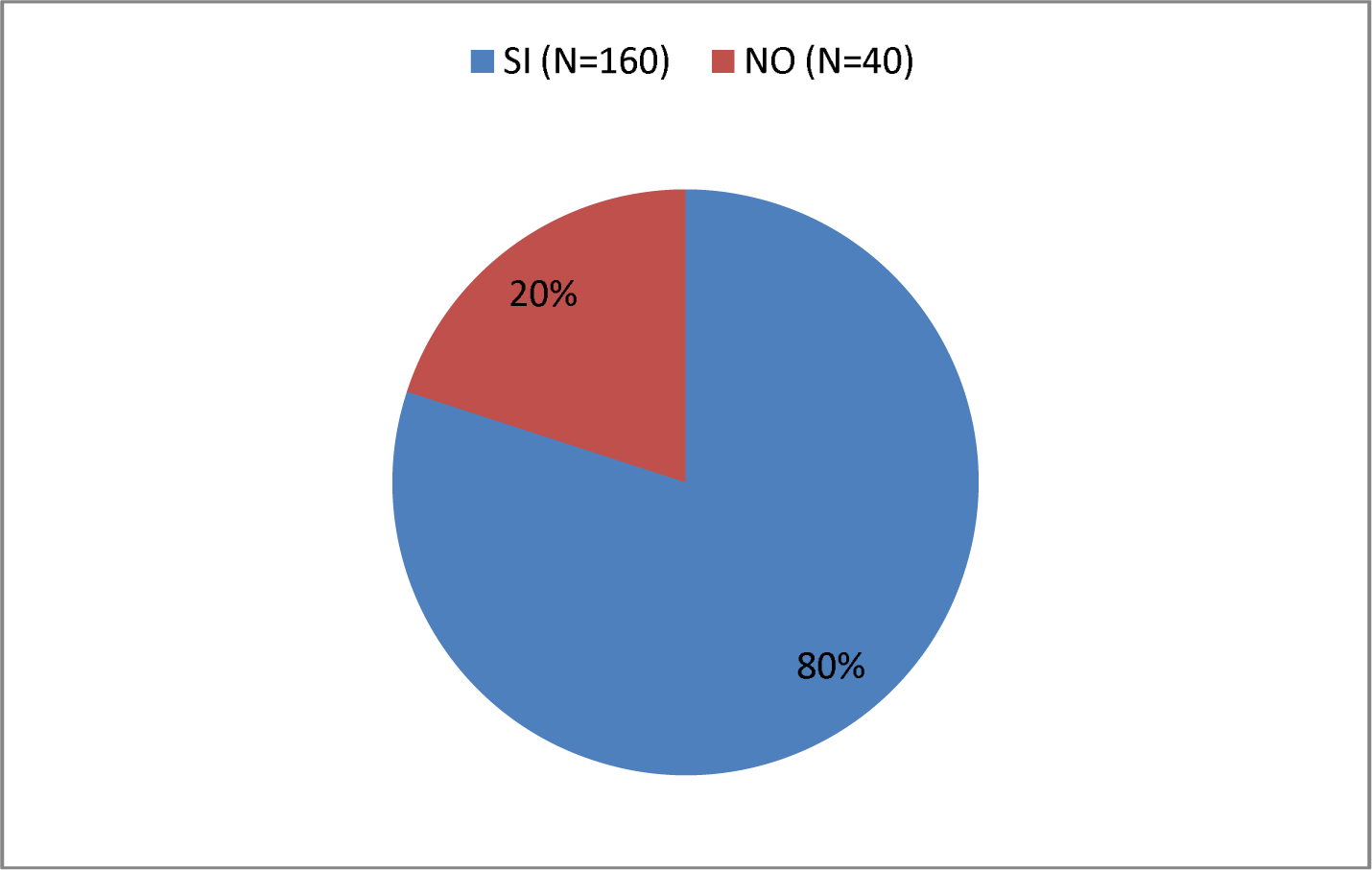
Do you know that the disease transmitted by the vinchuca affects the heart?

In this graph (Graph 30) you can see how of those interviewed (n=200) 42% (n=184) answered Yes to this question while 58% (n=116) answered No to it.

**Graph 30.**
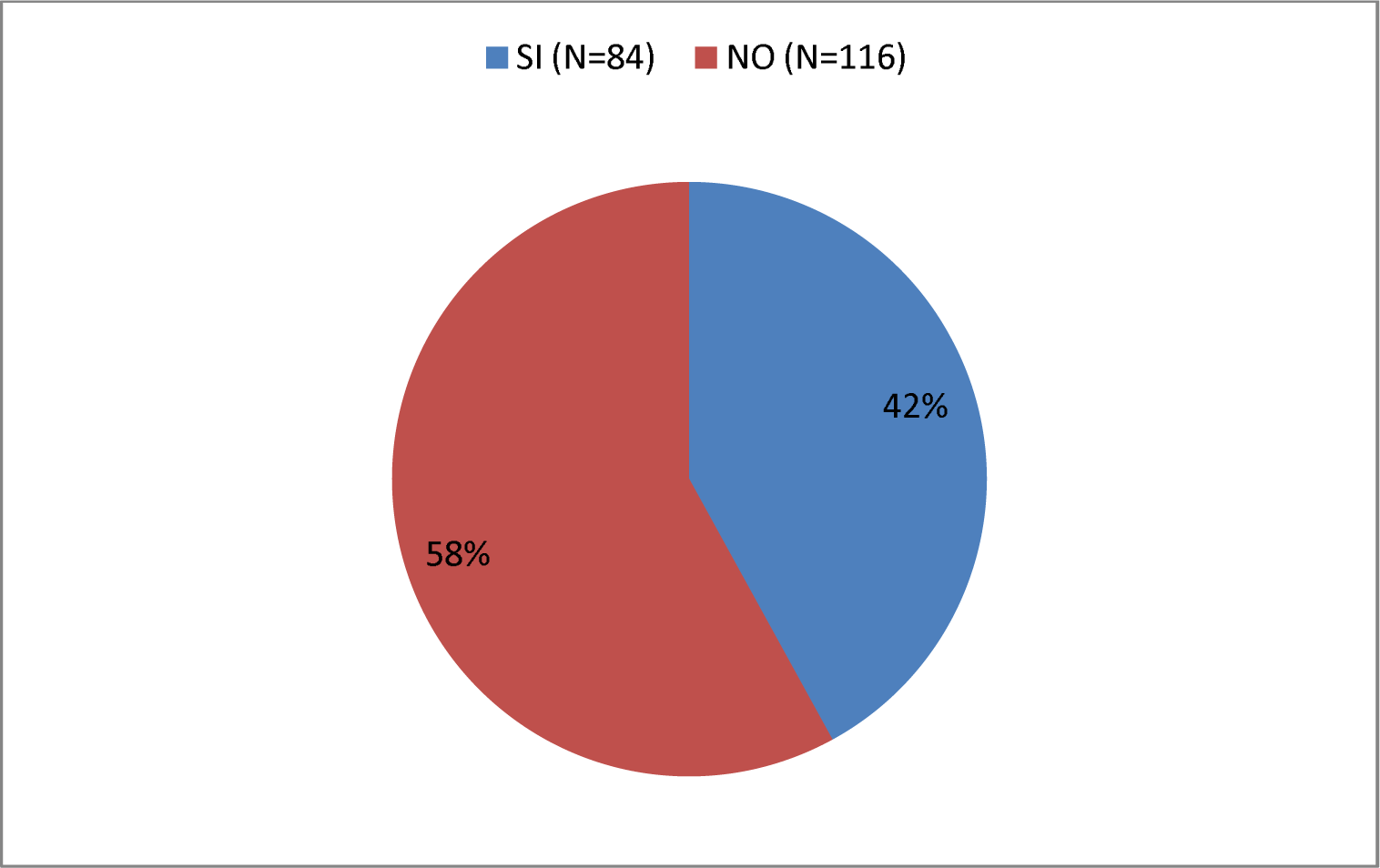
Do you know that the majority of cases of the disease transmitted by vinchuca do not have Chagas?

In the following graph (Graph 31) you can see how of those interviewed (n=200) 47% (n=95) answered Yes to this question while 53% (n=105) answered No to the same

**Graph 31.**
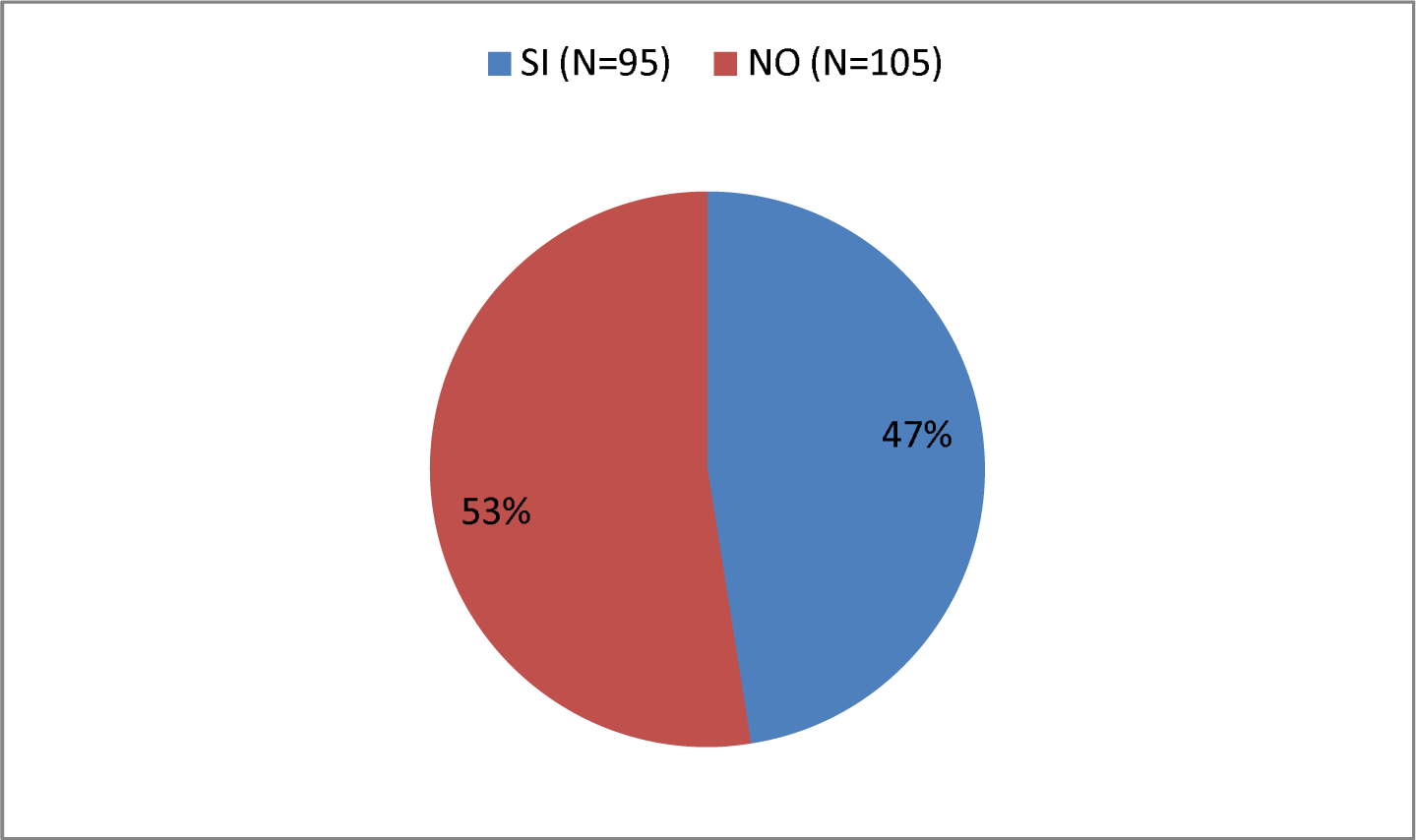
Do you know that vinchuca transmits the disease through its feces?

In this graph (Graph 32) you can see how of those interviewed (n=200) 51% (n=101) answered Yes to this question while 49% (n=99) answered No to it.

**Graf 32.**
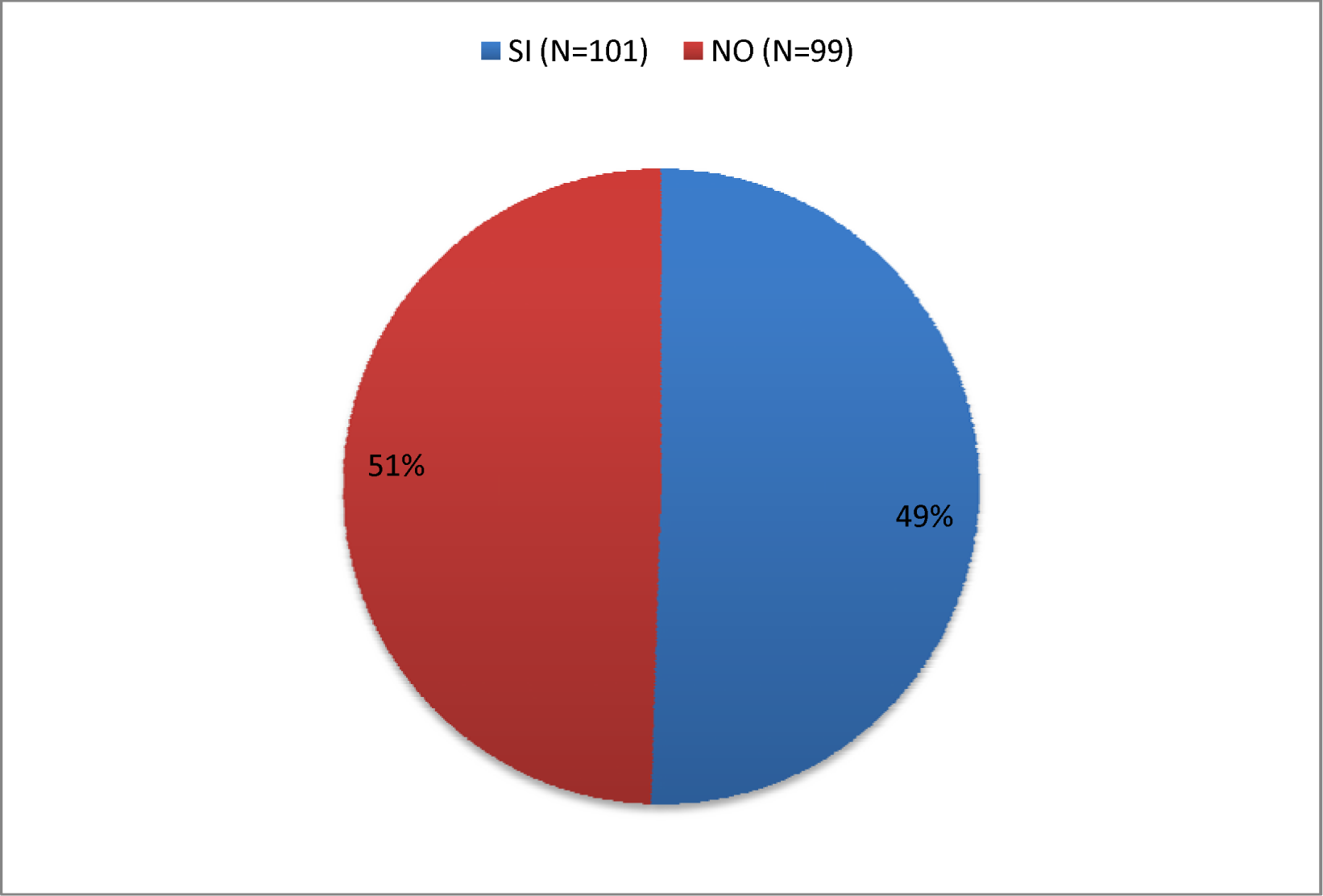
Do you know that vector transmission is carried out through wounds or skin abrasions caused by the vinchuca bite or through its eyes?

Regarding whether they knew that there are other ways of transmitting Chagas disease, the graph (Graph 33) shows that of the total interviewees (n=200) 13% (n=27) answered Yes to this question while a 87% (n=173) responded No to it.

**Graf 33.**
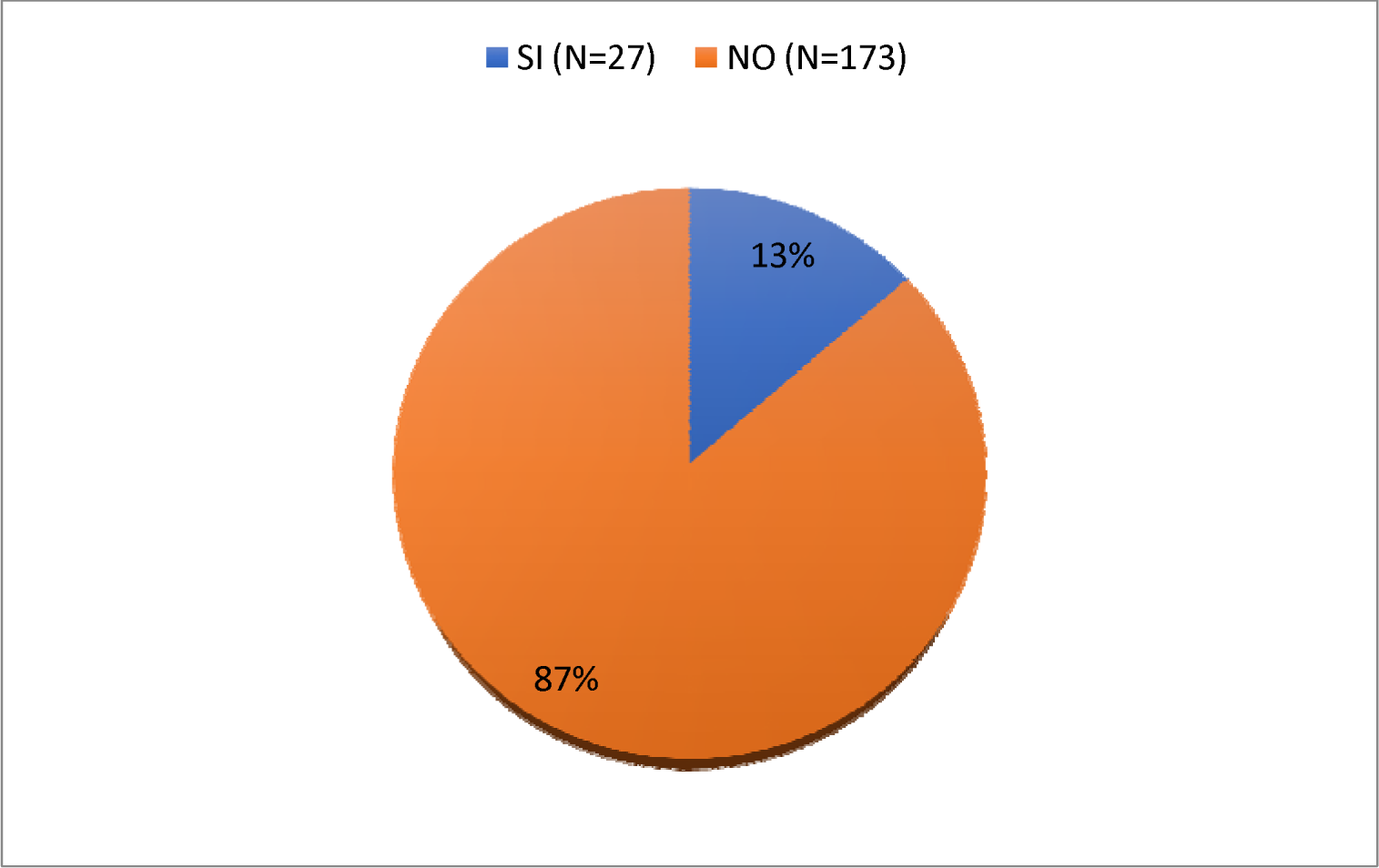
Do you know that there are other ways of transmitting Chagas disease?

In this graph (Graph 34) it can be seen that of the interviewees who knew another route of transmission or were aware of its existence (n=25), 56% (n=14) knew the congenital/vertical route of infection while that the remaining 44% (n=11) knew the route of transmission through blood transfusionthis question

**Graf. 34.**
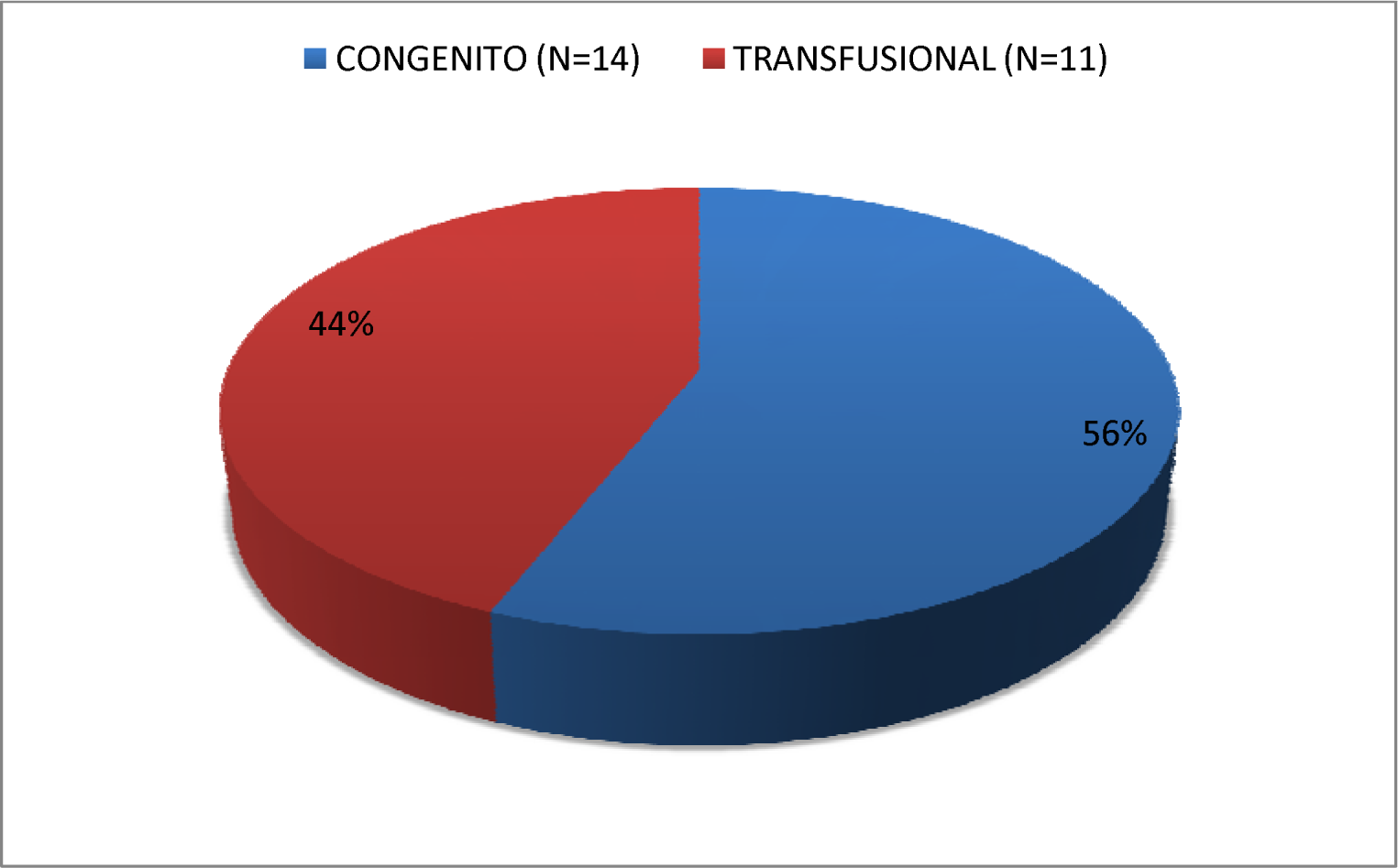
Which ones?

In this graph (Graph 35) you can see how of those interviewed (n=200) 10% (n=21) answered Yes to this question while 90% (n=179) answered No to it.

**Graf 35.**
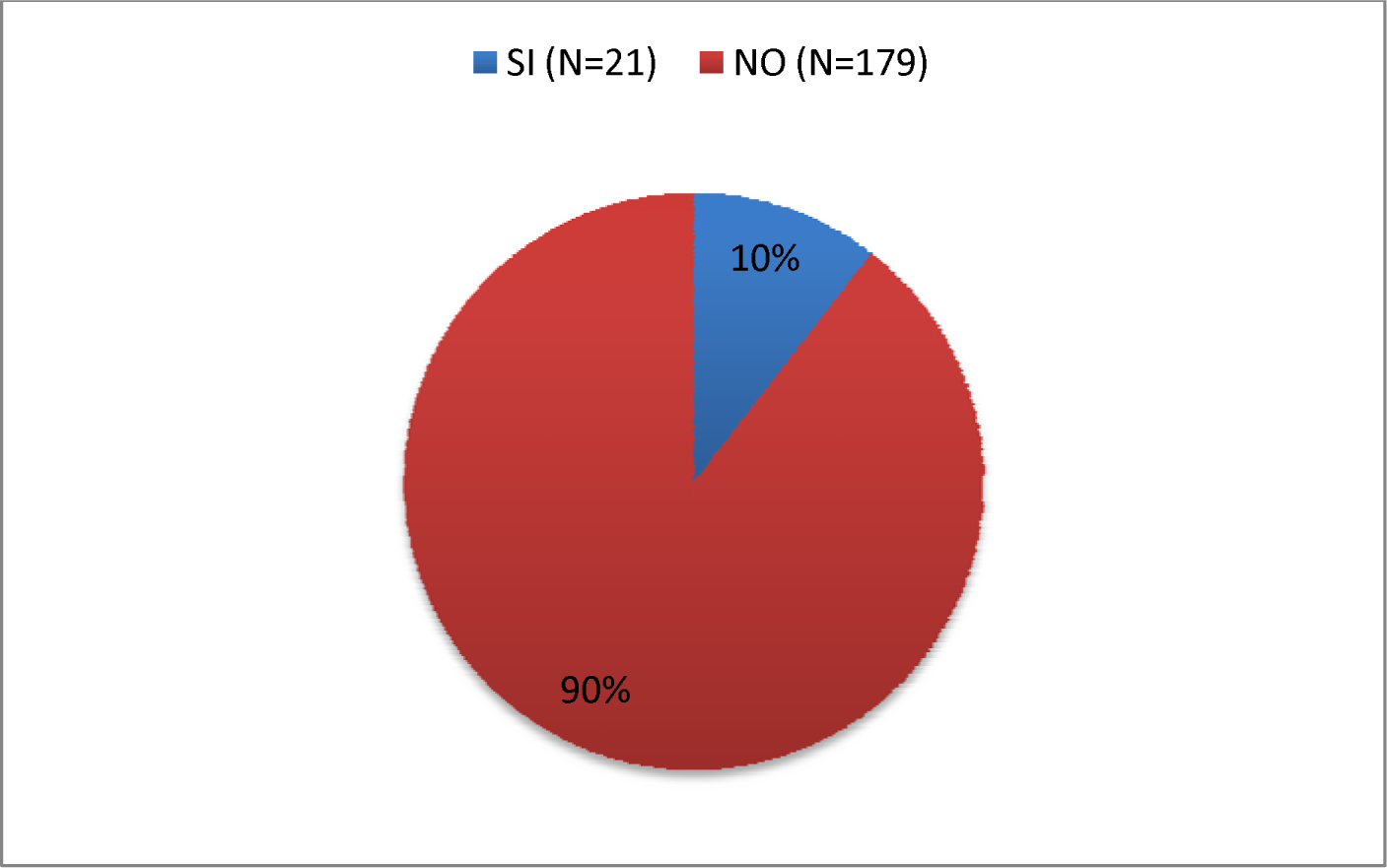
Do you know that vinchucas after death are important for the transmission of the disease?

A summary of the results obtained in this section of the survey can be seen in the following table:

**Table 1.** Optimun leve lof knledge.

| Question | Respuesta afirmativas |  |
| --- | --- | --- |
|  | Absolut frequency | Relative frequency |
| Recognize adult vinchucas | 68% | 82% |
| Recognize the presence of vinchucas feces | 41% | 41% |
| Know that the vinchuca's refuges are in the wall, ceiling, under the bed and cracks | 81% | 81% |
| You know that ranch-type homes favor the presence of vinchuca | 95% | 95% |
| Do you know that vinchucas transmit a disease? | 89% | 90% |
| You know that the disease transmitted by the vinchuca affects the heart | 80% | 80% |
| You know that the vinchuca transmits the disease through its feces | 47% | 48% |
| You know that vector transmission is carried out through wounds or abrasions on the skin or through the eyes | 51% | 51% |
| Do you know that there are other ways of transmitting Chagas disease? | 13% | 14% |
| Importance of dead vinchucas for transmission | 10% | 11% |

The following graph (Graph 37) represents the distribution of individuals (%) according to the number of affirmative responses in the survey regarding the level of knowledge about Chagas disease and Trypanosoma cruzi infection. To define the level of knowledge of each participant, it is stratified into: low (up to 3 correct questions, 25th percentile), intermediate (5 to 8 correct questions) and high (9 to 10 correct questions, 75th percentile).

Attributing to the NOC a level of knowledge of 100% (10 questions answered correctly), it is observed that, in the population sample studied, 12% have a low level of knowledge, 66% have an intermediate level, and only 22% have a level of knowledge. considered as high.

**Graph 37.**
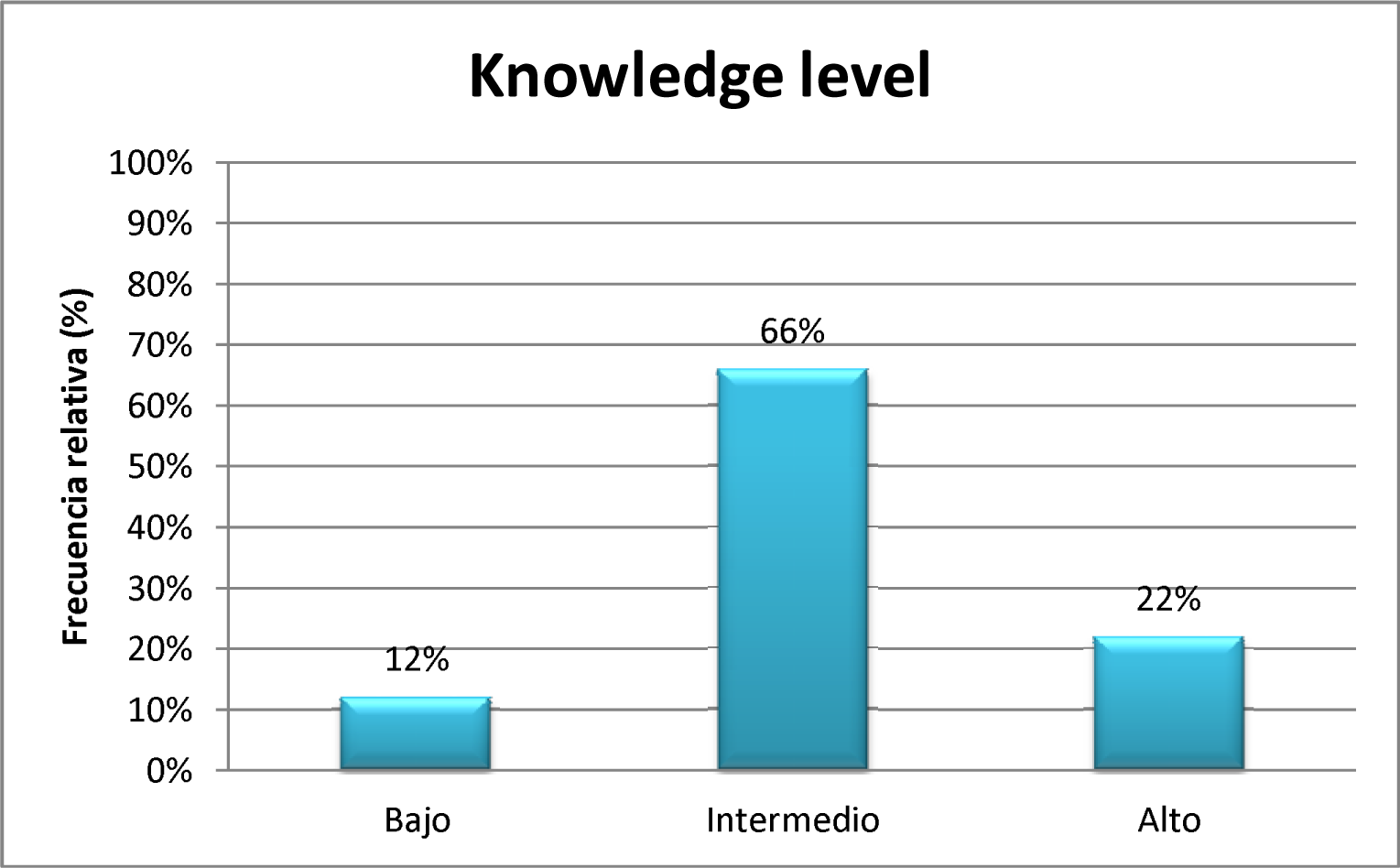
Distribution of the individuals studied (%) according to the level of knowledge of the population sample about Chagas disease and its vector. Low level of knowledge = up to 3 correct answers (25th percentile), intermediate = between 4 and 8; high = 9 or higher (75th percentile).

**Results of the third section surveyed: MEDICAL DATA**

**Graph 37.**
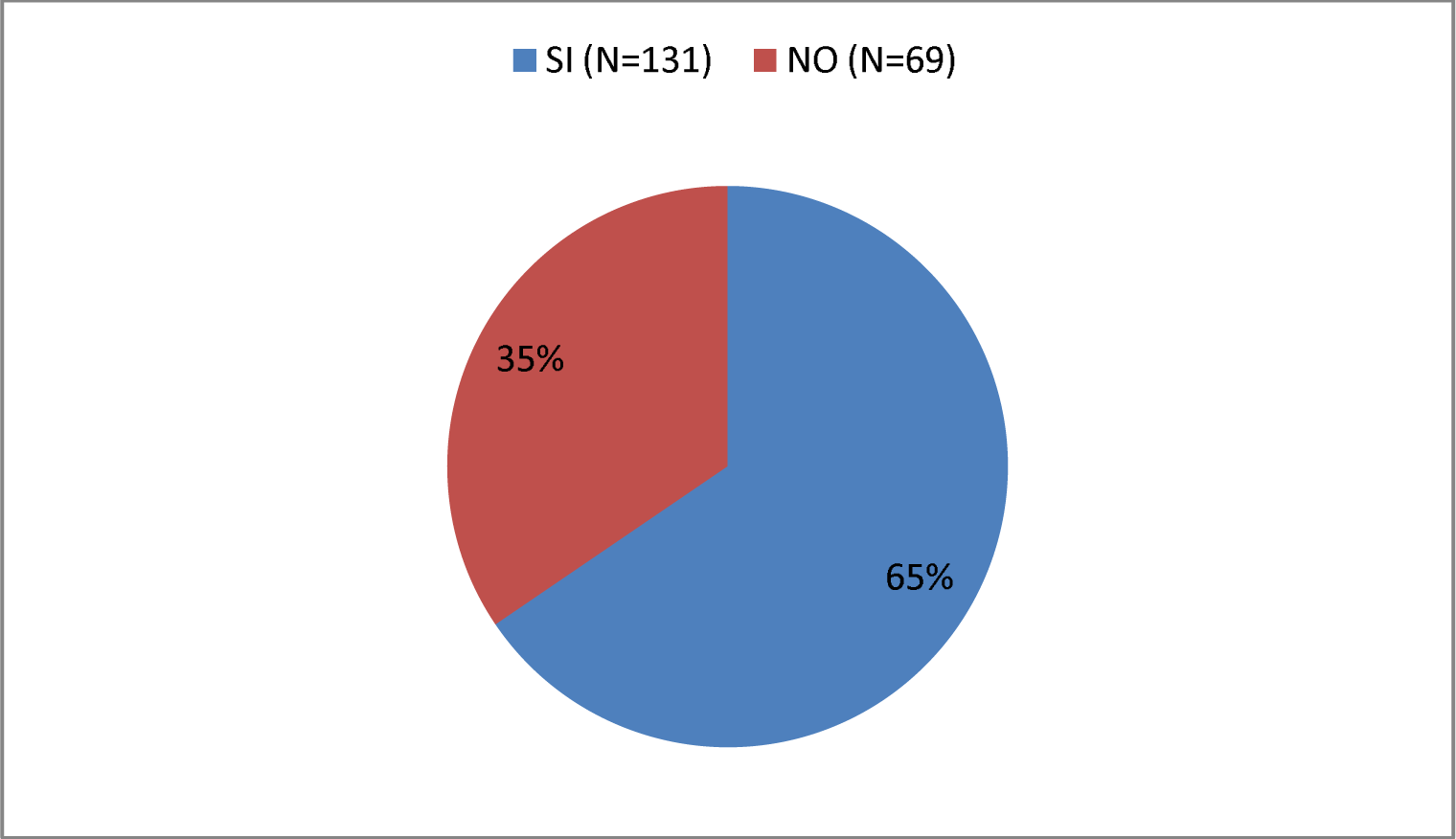
Do you carry out regular health check-ups with your doctor?--A

In this graph (Graph 38) you can see how of those interviewed (n=200) 31% (n=63) responded that they suffer from some relevant disease (HTN, arrhythmias, heart attack, heart failure, DBT, COPD, stroke, neoplasia, recurrent infections, immunosuppression), while 69% (n=137) answered no.

**Graf 38.**
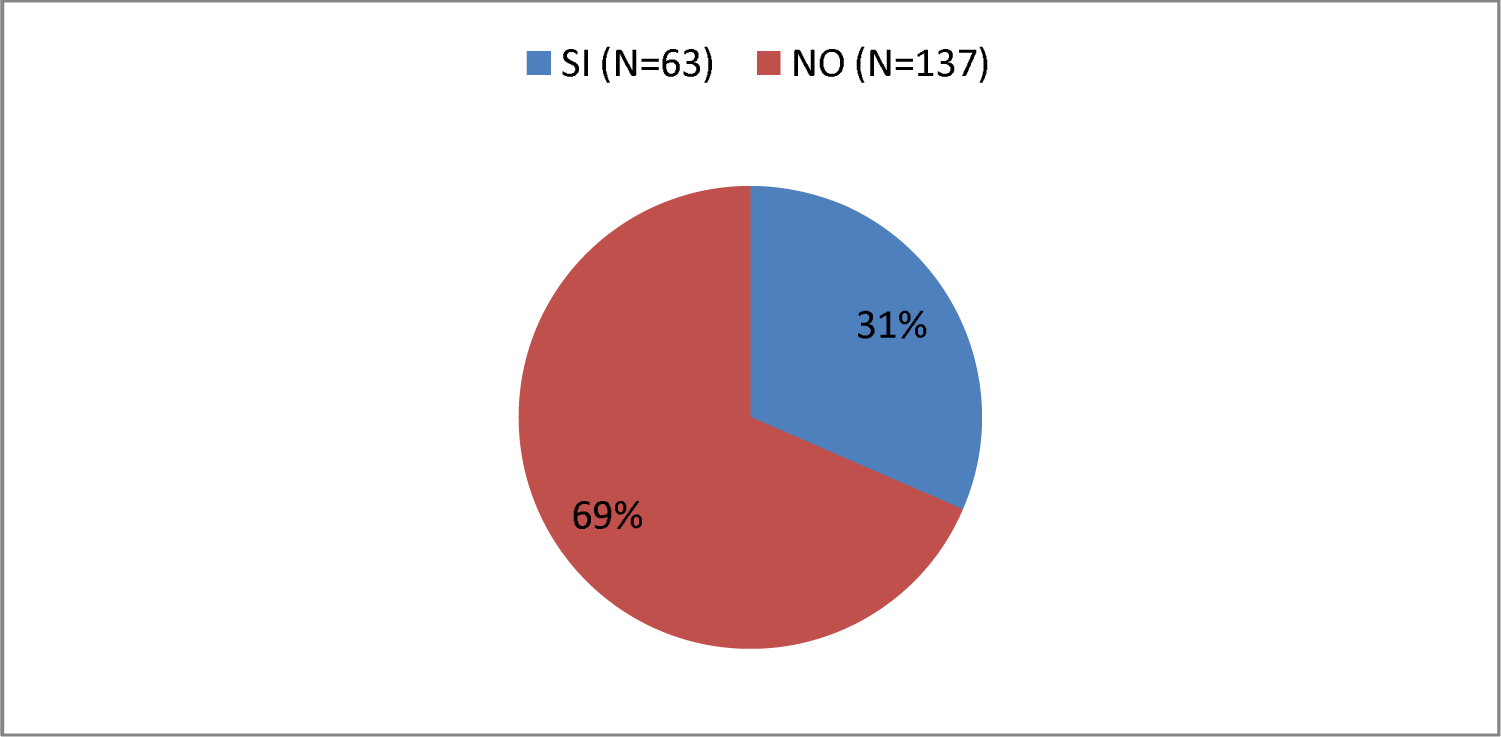
Do you suffer from any relevant illness?

Below you can see how 76% (n = 152) of all respondents underwent Chagas testing at some point in their lives, while 24% (n = 48) did not (Graph 39). Of the former, 24% (n = 36) had a positive Chagas analysis, while 76% (n = 117) were negative (Graph 40).

**Graph 39.**
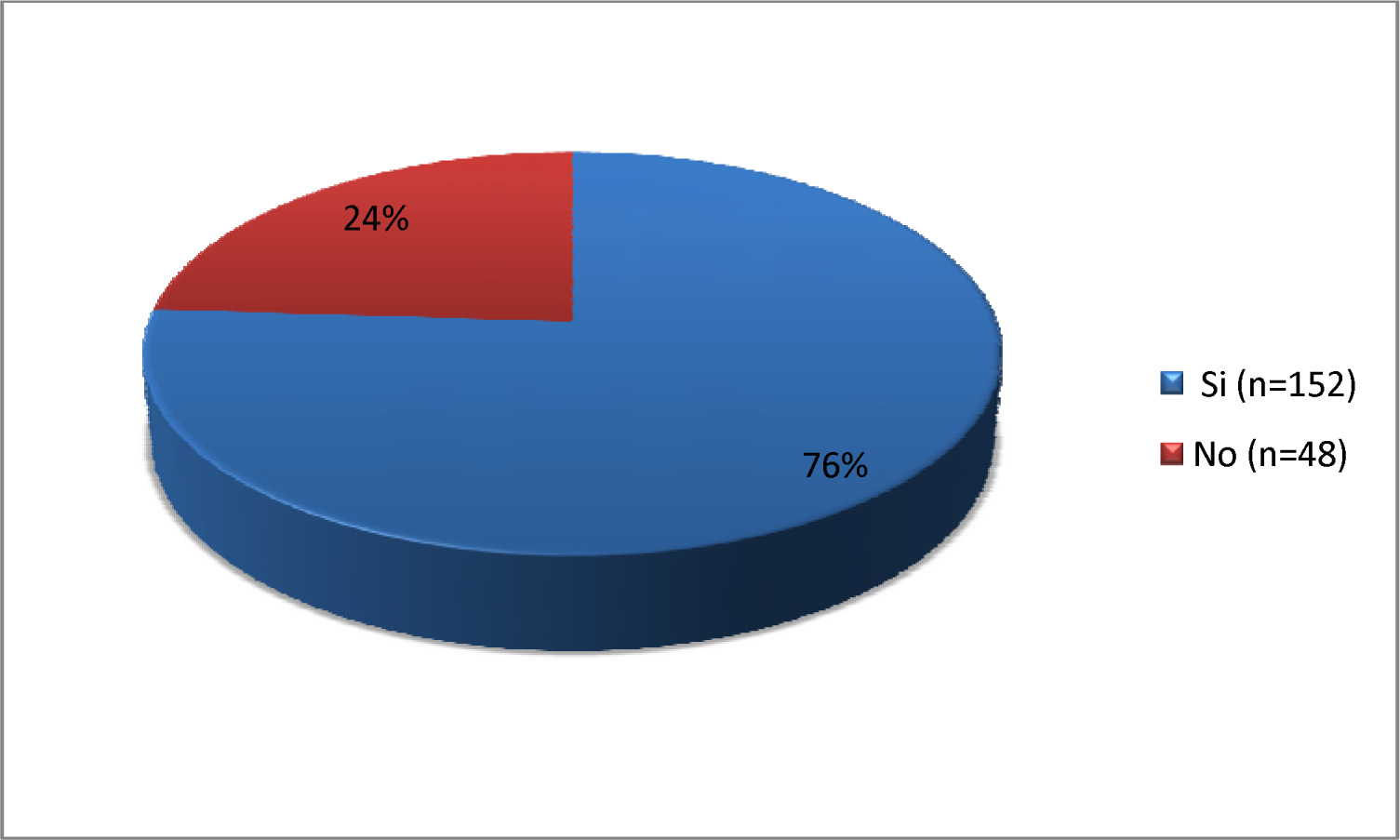
Have you ever had Chagas analysis in your life?

**Graph. 40.**
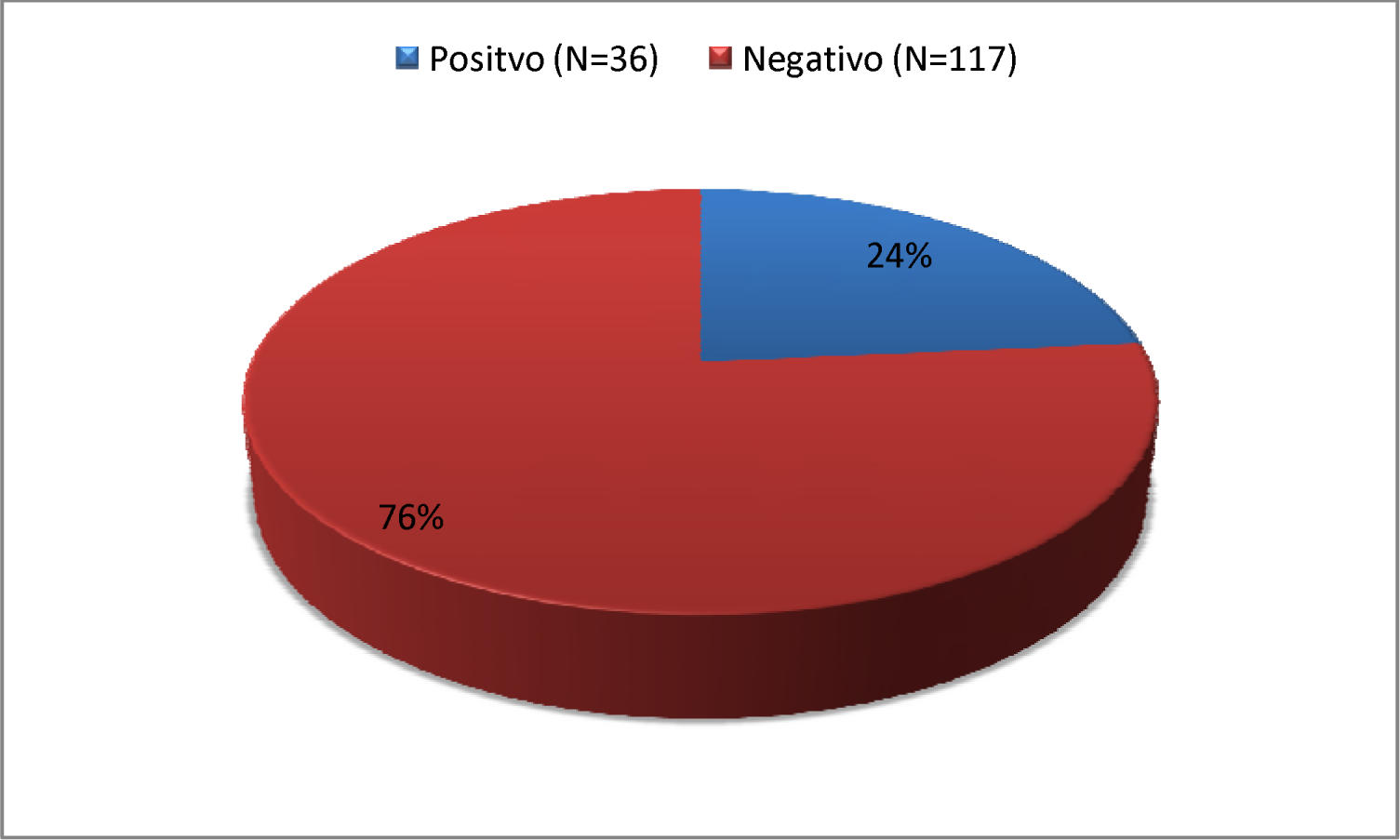
If the Chagas analysis has been done. Was it positive?

In the following graph (Graph 41) you can see how 56% (n = 20) of those surveyed went to the cardiologist this year, while 44% did not (n = 16).

**Graf. 41.**
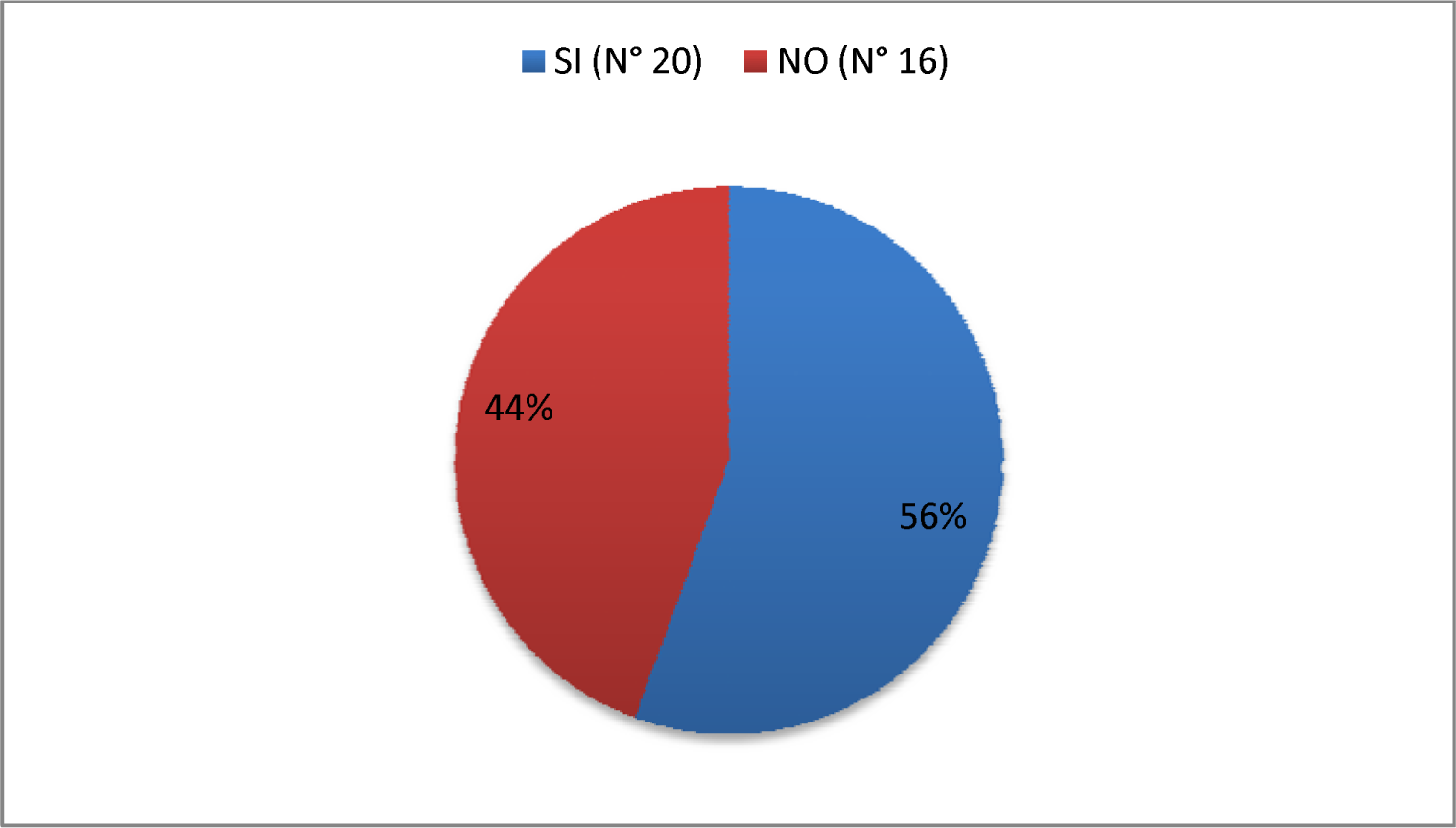
Have you been to the cardiologist in the last year?

Of the total number of respondents, 67% (n = 24) had an ECG performed in the last year, while 33% did not (n = 12) (Graph 42)

**Graf. 42.**
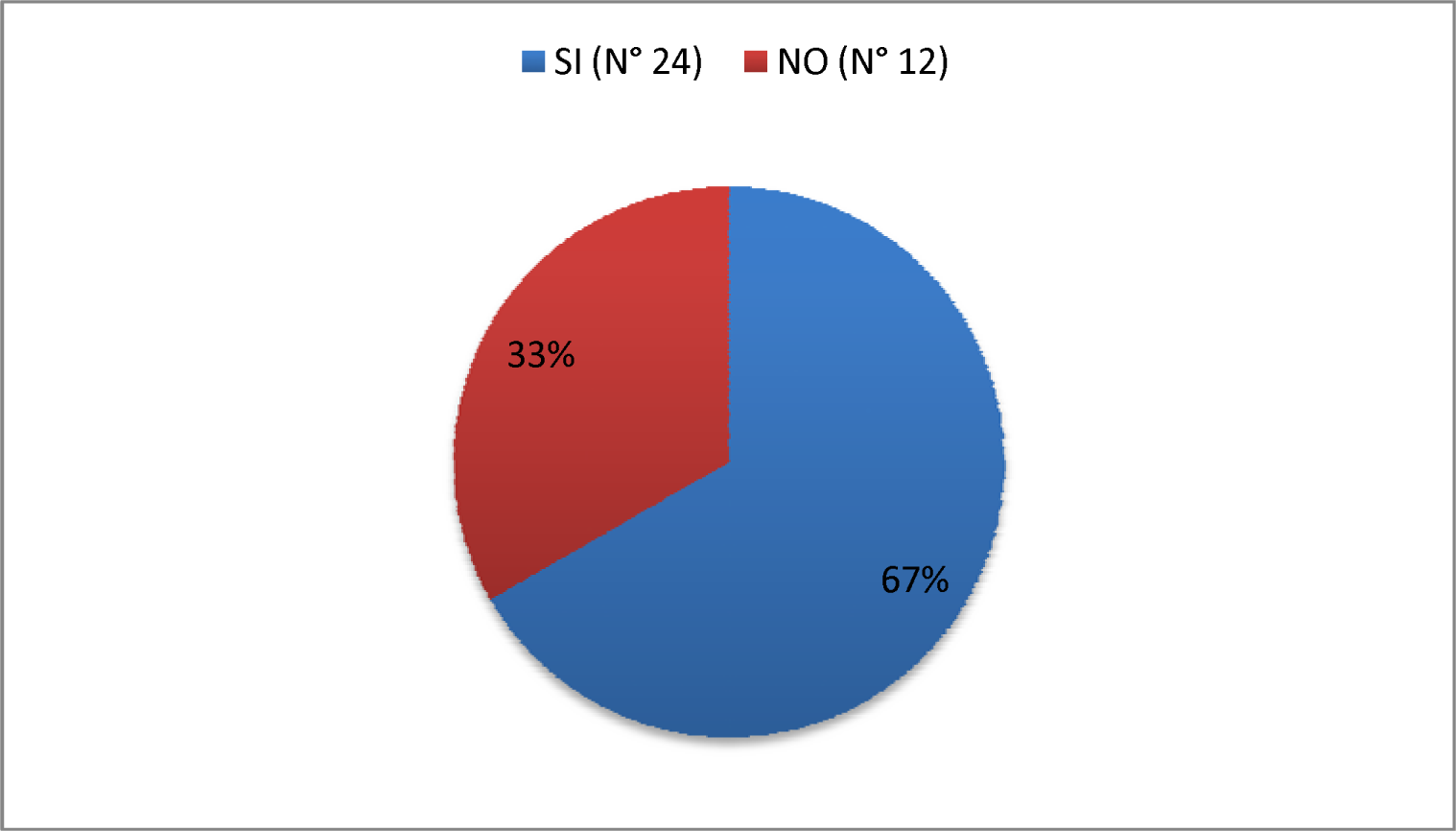
Have you had an electrocardiogram in the last year?

Graph 43 and 44 shows that 72% (n = 26) of those surveyed had symptoms or heart disease at some point during the last year, the most frequent symptoms being: feeling of shortness of breath and palpitations (30% each). one), chest pain (19%), fatigue (17%) and murmur detected (4%), respectively. On the other hand, 28% (n = 10) claim to have not had any heart symptoms/disease in the last year.

**Graf. 43.**
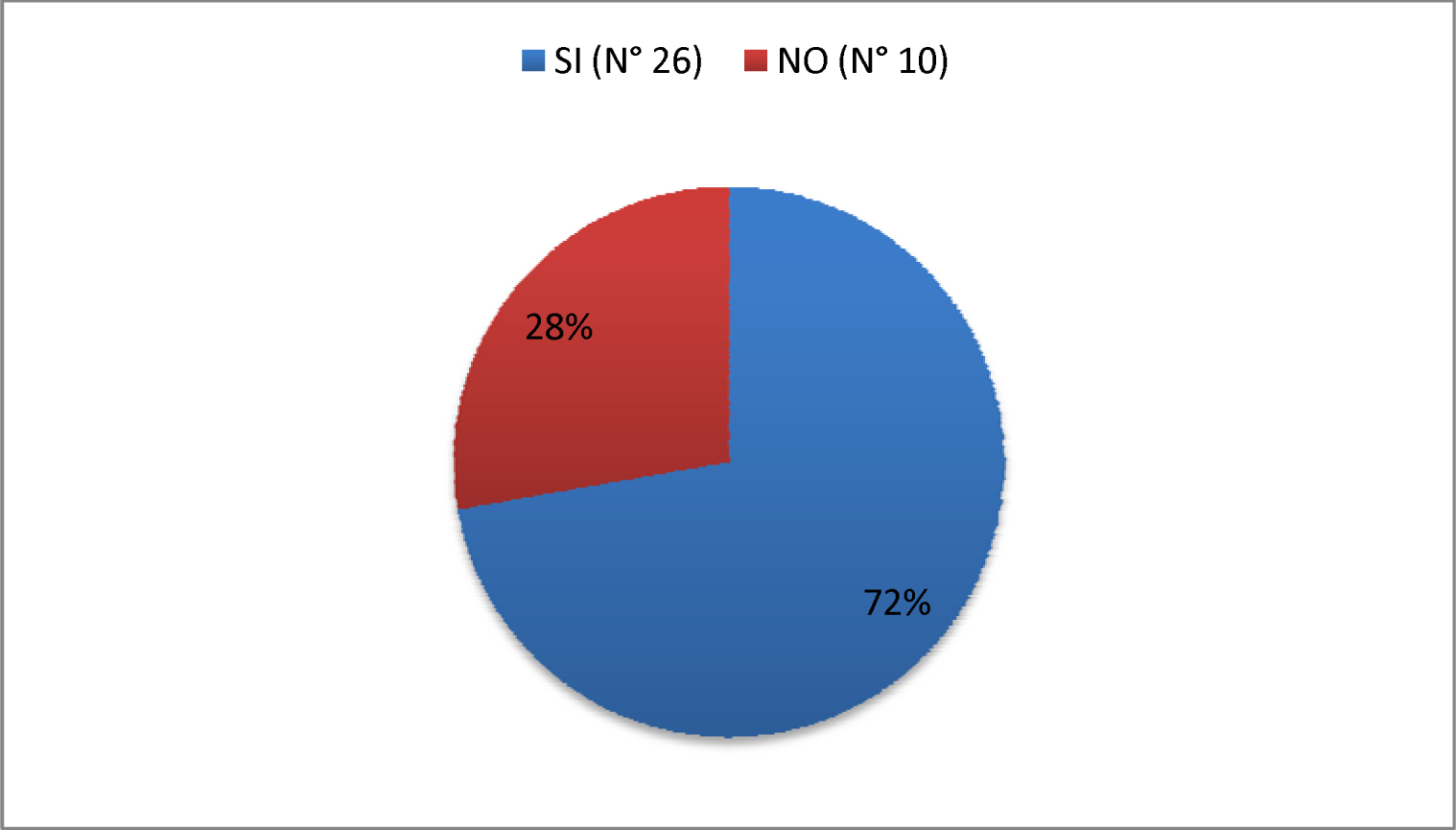
Did you have any heart symptoms/disease in the last year?

**Graf. 44.**
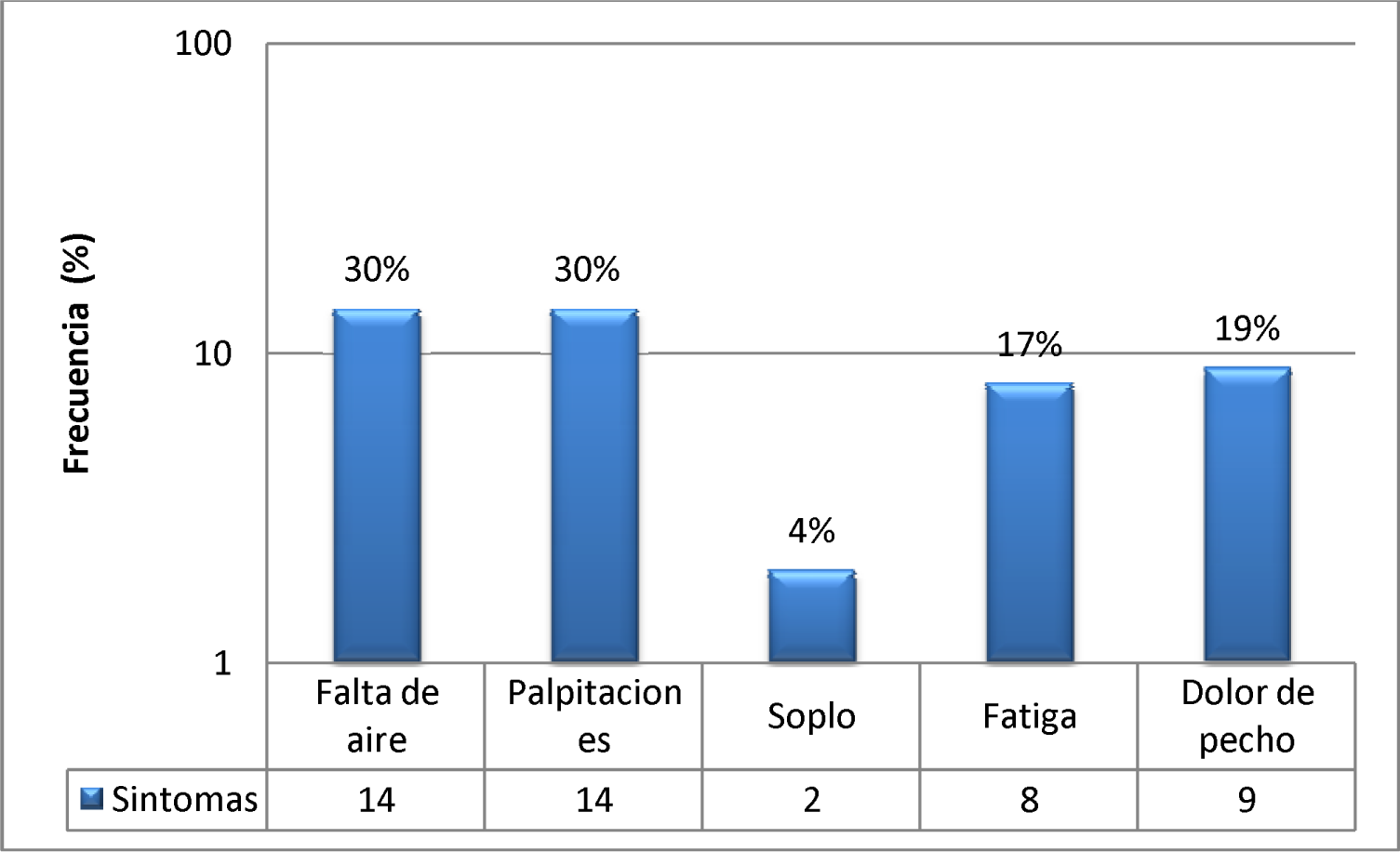
Most frequent symptoms

The following graph (Graph 45) shows how 83% (n = 30) do not and have not undergone any type of treatment for the disease, while 17% (n = 6) do.

**Graf. 45.**
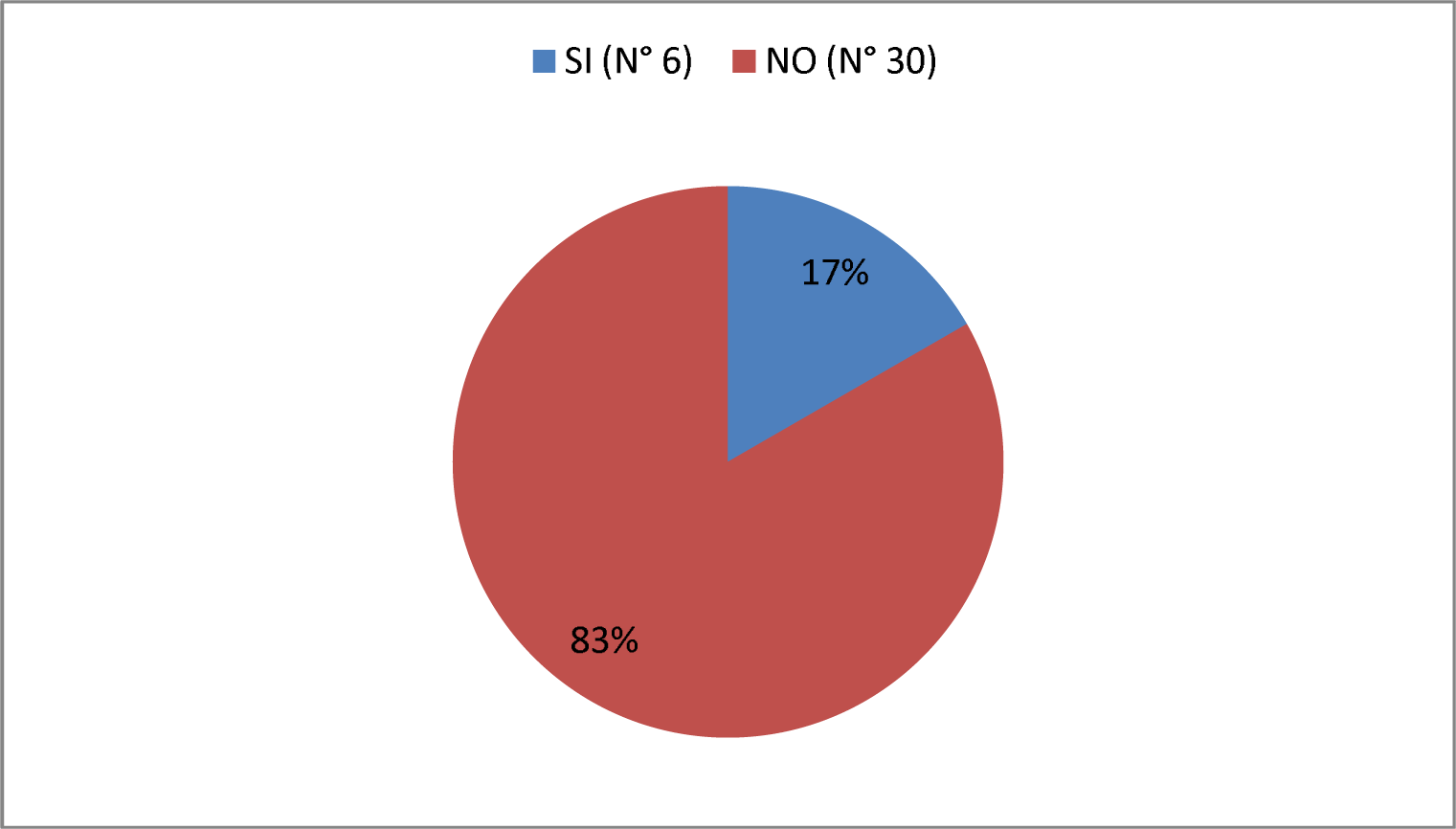
Do you carry out any type of treatment for Chagas Disease?

Once each of the questions-answers of the survey carried out on all patients was described (see Results section), we continued analyzing the statistical significance between the responses that correspond to the table of optimal notions of knowledge (Table 1) of the patients. CHAGASIC vs NON-CHAGASIC patients. To do this, we use the Chi square/Chi2 statistical test or also called Pearson distribution.

Within these questions we found that in only three of them there was a statistically significant difference (p < 0.05).

The first of them corresponds to the recognition by patients of feces or remains of vinchuca on the walls. We saw that the affirmative responses by non-chagasic patients was 37%, while in chagasic patients it was 55% (Chi = 3.845, p = 0.05)

The second was the knowledge that Chagas disease affects the heart. Non-chagasic patients answered affirmatively in 77%, while in chagasic patients it was 94% (Chi = 6.2, p = 0.01). In this question, the greatest statistically significant difference was observed between one group and the other.

The last question in which there was a statistically significant difference was in the question that refers to whether the respondents knew that the vinchuca transmits the disease through feces. In the non-chagasic group, 44% answered yes, while 64% of the chagasic patients answered yes (Chi = 4.728, p = 0.02).

Regarding the remaining 7 questions that make up the elements of optimal notions of knowledge (NOC Table), there were no statistically significant differences between non-chagasic and chagasic patients (Chi < 3.8, p > 0.05).

## Discussion

In the present work, the aim was to carry out a study on the level of knowledge about Chagas Disease and evaluate the possible risk factors in a population sample from the city of Añatuya, Santiago del Estero, Argentina, an endemic area of the disease. To this end, 200 surveys were carried out on people over 18 years of age who attended rural and regional health posts, and the Añatuya Zonal Hospital.

After analyzing the data, a slight predominance of females over males is noted (67% and 33%, respectively), with an average age of 30.4 years. Within these, there is a clear tendency towards having only completed primary studies or not having completed any studies, with the smallest number having completed secondary and tertiary studies. These data guide us to try to act with the corresponding educational tools such as providing classes, carrying out prevention and health promotion actions, and providing teaching materials for teaching about Chagas disease and its vector in primary schools and extracurricular, since most people fall into these last two groups.

It is important to remember that Chagas disease as a nest parasite is conditioned by various geographical factors, which include flora and fauna. Corresponding to this, there is a high frequency of the presence of animals in the home/peridomicile, mainly dogs, cats and chickens. According to the literature used, dogs represent reservoirs for T. cruzi infection, and their presence in the home is strongly correlated with a greater number of infected triatomines and a greater prevalence of Chagas Disease, mainly in children. As for chickens, they are a food source for triatomines and their presence impacts the domestic ecology of Chagas disease, since they increase the infestation of the home, leading to an increase in contact of the vector with domestic reservoirs. Within this analyzed panorama, it must be added that the average number of cohabitants per household was 6 people.

Regarding health coverage, the vast majority of those interviewed did not have health coverage, most of them receiving private care or in the hospital and paying the corresponding fees in the case of private services. This translates into a lack of health care and an overload of the public health system.

We found great variability in the place of residence of the people interviewed. This is a strength of the study, given that the aim was to search for a heterogeneous population in order to carry out a survey without bias by area where interviews were carried out, covering both rural and urban areas. Within these, the vast majority of people stated that they do not live with people who suffer from the disease.

The vinchucas easily colonize man’s homes, the rustic construction of the walls without plaster and the mud that compose them generate cracks and galleries, excellent shelter for the insect. The cracks, when covered by papers, leaves, cloths and pieces of rustic wood, generate new breeding sites for vinchucas. Regarding housing, this work shows that a little more than 50% had material housing with electricity and drinking water available, and 22% of the total respondents live in ranch-type houses. Of these homes, almost all have risky roofs, and the vast majority have risky walls, as well as having chicken coops, dogs or birds in their home or peridomicile. Although the majority of people stay in material housing, the fact that ranches continue to exist and in large numbers reflects that the actions to eradicate ranches are insufficient. We also observed that the number of people who do not have waste collection service is greater, so they must do it on their own. This information is worrying since, without correct garbage collection, disorder is encouraged, an environment conducive to vinchuca breeding sites.

Approximately half of the population surveyed stated that they had ever witnessed a vinchuca or traces (feces, molts or eggs) in their home or in the peri-domicile, although the majority received fumigation services to combat the vector of the disease, with a frequency of 1 fumigation per year as the predominant value. This data obtained should encourage us to continue with tests and research regarding the effectiveness of insecticides generally used during spraying of homes, monitoring of risk homes post-fumigation and their relationship with the resistance of vinchucas to these substances. (14)

Previous studies have recognized the importance of inhabitants of endemic areas knowing their situation perfectly in order to protect themselves and their families. In relation to this, in the present work a general lack of basic knowledge of the disease was identified, mainly in five aspects: the recognition of the feces of the vinchucas present on the walls, that the majority of the vinchucas do not transmit Chagas, that The disease is transmitted through their feces, other routes of transmission other than vector transmission and that the vinchucas after death continue to be important for the transmission of the disease. This knowledge was not present in more than half of the respondents, which indicates a level much lower than optimal in an endemic area such as the city of Añatuya. This level of general ignorance may be due to the large number of people who did not complete their primary education or to a deficit in public education. Although this issue was not fully addressed, it is a debt of public schools to provide information to students about the disease, especially in an endemic area.

Regarding knowledge of the disease, most recognize the adult vinchuca, but not the presence of feces on the walls. It is proposed to superimpose in subsequent studies whether the observation of vinchucas in homes and surroundings is correlated with the ability to recognize adult vinchucas in order to avoid simple errors by the interviewees. The interviewees presented greater knowledge in 3 aspects: that vinchucas bite humans, that ranch-type housing favors the presence of vinchucas, and that they feed on blood.

It should also be taken into account that many of the respondents are located at considerable distances from the study centers (especially those who live in ranch-type homes) and that the majority of them do not have the possibility of accessing this information., which presumes a failure in the dissemination of information outside educational barriers. It should not be forgotten that the social dimension of the disease is fundamental, since many inhabitants of rural areas do not want to abandon their ranches due to a feeling of “uprootedness”, rejecting the offer of social housing.

Finally, when analyzing and comparing Chagas patients with non-Chagas patients, there is a difference in knowledge in favor of Chagas patients in three important points: the recognition of the feces of the vinchucas on the walls, in which the disease affects the heart and that the vinchuca transmits the disease through its feces. Taking into account that 30% of chronic Chagas sufferers show symptoms of complications of the disease, we believe that this motivates them to enter the health system to consult and, in this way, they usually receive more information and from a better source. It should be remembered that heart disease is the most common complication of the disease in our country, which can make an important difference in the aforementioned question. Of all the chagas patients, the symptoms for which they most consulted were the feeling of lack of air (dyspnea) and palpitations. On the other hand, regarding chagas patients, we noticed that 83% of those surveyed do not undergo any treatment.

### Strengths of the study

- Large N (>100), which helps reduce random error. And normal distribution is established according to the central limit theorem.
- We interview at different times and places, so there is a greater probability of obtaining a sample that is not biased by time or place of attendance. Both rural and urban population; population surveyed inside and outside the hospital.
- In order to avoid reporting bias, time is taken to explain the question to each interviewee and for them to understand it, in the most objective way possible.
- We avoid observation bias since at the end of the survey we ask whether or not you have a diagnosis of Chagas disease. From this, we evaluate our results on knowledge of the disease.
- Selective participation or selection bias:
• To avoid this we emphasized that the information was going to be used for surveys and that it would help to better understand the disease.

### Limitations of the study

- Biases inherent to the type of study:
• Inability to demonstrate causality due to lack of temporal sequence.
• It does not allow reporting on a real association between variables, the most they allow is reporting on the relationship or that there appears to be an association.
- Confusers
• Associate the survey with the vector without paying attention to other forms of transmission (transfusion, congenital). It was not asked how the chagas was acquired.

## Conclusion

The results obtained during the present work carried out in the City of Añatuya, Santiago del Estero, mark a level of knowledge about the disease that is below optimal. According to the list of elementary notions that constitute the NOC of Chagas disease, 66% of the population has an intermediate level of knowledge and only 22% has a high level of knowledge, close to the optimal level for an endemic region. On the other hand, the level of knowledge was similar between chagasic and non-chagasic patients, therefore being chagasic does not determine having a greater degree of information about the disease.

Furthermore, a good part of the surveyed population has risk factors for the presence of vinchuca, accentuating the persistence of this endemic disease.

The socio-cultural and demographic conditions of the population are related to the risk factors for Chagas Disease. In most of our sample, socio-cultural and demographic conditions are present that are risk factors for Chagas Disease. Among these, the presence of risky housing, disorder/dirt, and the large percentage of domestic animals and animals stand out. non-domestic in the home or peridomicile, being facilitators for the presence of vinchuca and therefore Chagas Disease.

We were also able to promote health education by carrying out surveys since, once the survey was completed, we explained the theoretically correct answers, so that the population would have greater knowledge of the disease and thus be able to prevent it.

We believe that the fight against ignorance about the risk factors for Chagas disease would imply an important advance on this disease, leading the inhabitants of endemic areas to a better understanding of their reality and the acquisition of habits that allow them to be the protagonists. of your own well-being.

Finally, we consider it essential that the population that lives exposed to the risk of contracting Chagas disease has the necessary knowledge to be able to fight it through their daily actions. It is necessary that this knowledge be promoted not only by health agents but also that it must be incorporated into the educational system and the community system so that transmission and learning by the entire population is guaranteed.

### Suggestions and proposals

- Carry out community awareness campaigns about the disease, emphasizing basic information to avoid its spread by the vector, and the risk factors that can favor the proliferation of the vinchuca. This can be carried out in public places (squares, parks, libraries, clubs), in educational settings (primary and secondary schools), at municipal events and/or local celebrations. This would cover the busiest places in the city of Añatuya, including the majority of the population.
- Include mandatory talks/classes within the school teaching program of primary schools in endemic areas such as Santiago del Estero, in which basic information about Chagas is discussed, given by trained health personnel.
- Continue carrying out fumigation tasks in the neighborhoods most affected by vinchuca, those with the greatest number of infected people, ranches and/or homes with risk structures.
- Insist on carrying out the Ranch Housing Eradication Plan to promote the elimination of the vector.
- Provide annual update classes on the diagnosis and treatment of Chagas Disease to health professionals at the Añatuya Zonal Hospital. In this way, the active role of the doctor in seeking early detection of the disease would increase and all doctors would have the same information, received from a reliable source.
- Encourage serological studies to be carried out on all people who were born and/or lived in endemic areas (where there are vinchucas) or whose mothers were born or lived in these areas, especially children and women of childbearing age.
- Design an official database of all Chagas patients in the city to require periodic checks on them. In this way, controlled monitoring of the disease could be implemented, timely treatments of acute, congenital or vector-borne and chronic infection could be implemented, and thus be able to try to avoid complications of the disease, as well as quantify the benefit of the prevention measures implemented.

## Data Availability

All data produced in the present work are contained in the manuscript

## Notes

### Competing Interest Statement

The authors have declared no competing interest.

### Funding Statement

This study did not receive any funding

### Author Declarations

Ethics committee of the Interzonal Hospital of the city of Anatuya, Santiago del Estero, gave ethical approval for this work.

